# Phenotypic evolution of SARS-CoV-2: a statistical inference approach

**DOI:** 10.1101/2022.08.25.22279206

**Authors:** Wakinyan Benhamou, Sébastien Lion, Rémi Choquet, Sylvain Gandon

## Abstract

Since its emergence in late 2019, the SARS-CoV-2 virus has spread globally, causing the ongoing COVID-19 pandemic. In the fall of 2020, the Alpha variant (lineage B.1.1.7) was detected in England and spread rapidly, outcompeting the previous lineage. Yet, very little is known about the underlying modifications of the infection process that can explain this selective advantage. Here, we try to quantify how the Alpha variant differed from its predecessor on two phenotypic traits: the transmission rate and the duration of infectiousness. To this end, we analysed the joint epidemiological and evolutionary dynamics as a function of the Stringency Index, a measure of the amount of Non-Pharmaceutical Interventions. Assuming that these control measures reduce contact rates and transmission, we developed a two-step approach based on *SEIR* models and the analysis of a combination of epidemiological and evolutionary information. First, we quantify the link between Stringency Index and the reduction in viral transmission. Secondly, based on a novel theoretical derivation of the selection gradient in an *SEIR* model, we infer the phenotype of the Alpha variant from its frequency changes. We show that its selective advantage is more likely to result from a higher transmission than from a longer infectious period.

## 1 Introduction

In December 2019, acute pneumonias of as yet ‘*unknown etiology* ‘ were increasingly reported in Wuhan, the capital of the Hubei Province in Central China [30]. Since then, the infectious agent responsible of this emerging zoonosis, a virus of the family *Coronaviridae* named SARS-CoV-2 (Severe Acute Respiratory Syndrome-CoronaVirus-2), has spread worldwide, causing the pandemic COVID-19 (Coronavirus Disease-2019) [49] that is still ongoing today.

The possibility of a rapid SARS-CoV-2 adaptation was initially met with considerable scepticism [16, 40]. Indeed, compared to other single-stranded RNA viruses, the mutation rate of SARS-CoV-2 is relatively low (estimated at the onset of the pandemic around 6.8*−*9.8*×*10^*−*4^ substitution.site^*−*1^.year^*−*1^ [45, 46]). Besides, all the observed mutations in SARS-CoV-2 were initially thought to be neutral or slightly deleterious. The occasional rise of some mutations could be due to demographic stochasticity [15, 9] but the dramatic rise of specific mutations in different regions of the world challenged the hypothesis that none of these mutations were beneficial. In particular, the analysis of the emergence and the spread of several Variants of Concern (VOCs) across the world – e.g. Alpha (lineage B.1.1.7), Delta (lineage B.1.617.2) or Omicron (lineage B.1.1.529) (see for example CoVariants [21] or Nextsrain [17]) – demonstrated that these variants carry adaptive mutations that explain their faster rate of spread in the human population [31]. However, each of these mutations may act on various dimensions of the fitness landscape of the virus and affect different life-history traits. It is therefore much less clear *why* specific variants are favoured. In other words: which phenotypic trait(s) can explain this increase in viral fitness? Viral fitness is governed by multiple life-history traits like the transmission, the virulence or the recovery rates of the virus [9]. It is crucial to understand which traits are involved in the increase in fitness because they may have very different implications for epidemiological dynamics and public health. For instance, an increase in the transmission rate or in the duration of infectiousness both lead to an increase in viral fitness but they may have distinct consequences for the efficacy of Non-Pharmaceutical Interventions (NPIs), implemented to mitigate the epidemic. It is therefore very important to understand and track this adaptation to optimize our control strategies.

In the following, we will focus on the first of these VOCs: the lineage B.1.1.7, categorised as *Variant of Concern 202012/01* and afterwards named “*Alpha variant* “. This variant emerged in early fall 2020 in the South-East region of England [36, 47] and then spread rapidly across the country (**Fig. 1**). The reproduction number of the Alpha variant (i.e. its expected number of secondary infections) was estimated to be 40-100% higher than for the previous lineage [5, 47]. Several studies aimed to unravel what phenotypic differences could explain this increased fitness. First, Davies *et al*. [5] explored various underlying biological mechanisms and suggested that a higher transmission rate per contact for the Alpha variant was the most parsimonious explanation, but that a longer duration of infectiousness – merely increasing the number of opportunities of transmission – could also explain the data very well. Blanquart *et al*. [3] developed another methodological approach considering three phenotypic traits: the overall reproduction number, the mean and the standard deviation of the generation time distribution of the infection. They showed that the selective advantage of the Alpha variant was likely to be driven by a higher reproduction number with an unaltered mean generation time.

**Figure 1:**
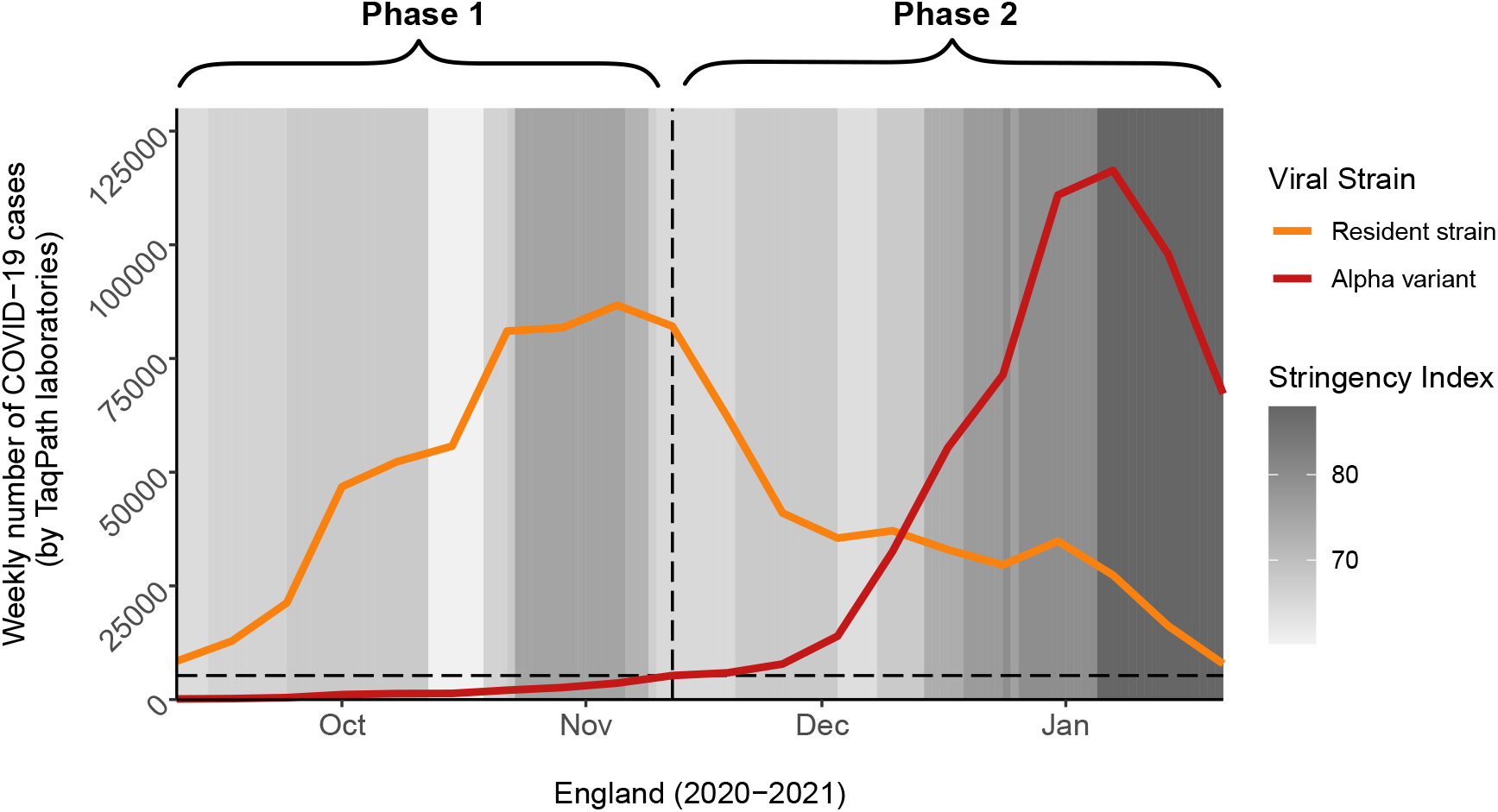
The two consecutive phases of the analysis of the spread of the Alpha variant. In phase 1 (before the emergence of the Alpha variant), we assume the epidemic is driven solely by the resident strain; in phase 2 (after the emergence of the Alpha variant), the epidemic results from the joint dynamics of the resident strain and the Alpha variant. In the first step of our analysis, we estimated the impact of the Stringency Index (a measure of the amount of NPIs implemented to mitigate the epidemic, from 0 (no control) to 100 (stringest control)) on the propagation of the resident strain during phase 1. In the second step of our analysis, knowing the impact of NPIs, we estimated the phenotypic differences between the resident strain and the Alpha variant during phase 2. The dates reported on the chart match the middle of each week (Thursday). We set the end of phase 1 when the Alpha variant reached 5% of the cases tested positive at the national scale (horizontal dashed line). For the sake of simplicity, we show data at the national scale but the starting date of phase 2 varied among regions (see **Fig. S1** and **Methods, §4.1**).

The present work is a new attempt to characterise the life-history traits of the Alpha variant, for which we consider two phenotypic traits: (i) the transmission rate and (ii) the recovery rate (inverse of the mean duration of infectiousness). We propose a novel approach to estimate these two phenotypic traits based on the analysis of the time-varying fluctuations of the selection coefficient driven by the variability in the intensity of NPIs used to limit the spread of the virus. As pointed out by Otto *et al*. [32], the selection coefficient of the Alpha variant (i.e. the slope of the change in its logit-frequency) varied with the intensity of NPIs, measured by the “*Stringency Index*”, a composite score published by the Oxford COVID-19 Government Response Tracker (OxCGRT) [18]. In [9] and [32], control measures that reduce contact rates between infectious and susceptible hosts are predicted to reduce the (relative) selective advantage of variants that have a higher transmission rate – in addition to slowing down the spread of the epidemic – but without affecting the selective advantage of variants that have a longer duration of infectiousness. In the following we exploit these contrasting effects of NPIs on the selection coefficient to infer the transmission rate and the mean duration of infectiousness of the new variant.

We use a stepwise approach of two consecutive phases of the epidemic (**Fig. 1**). First, we focus on the analysis of the epidemiological dynamics taking place just before the emergence of the Alpha variant (i.e. just before it reached 5% of the positive cases) and we infer the relationship between the Stringency Index and the effectiveness of the control measures (NPIs) on the viral propagation in the UK. Second, we derive a novel expression for the selection coefficient of a variant in a susceptibleexposed-infectious-recovered (*SEIR*) model. Knowing the impact of NPIs on the viral propagation from the first step, we use our expression of the selection coefficient to infer the effects of the mutations of the Alpha variant on (i) the transmission rate and (ii) the mean duration of infectiousness from the analysis of the evolutionary dynamics taking place, in each region of England, just after the emergence of the variant (i.e. just after it reached 10% of the positive cases).

## 2 Results

In the first step of the analysis, we develop an *SEIR* model (see equations (3) and **Fig. S3**) to capture the effect of the control measures *c*(*t*) on the epidemiological dynamics. The effectiveness of these control measures have been quantified and monitored with the Stringency Index [18]. As shown in the methods (§4.1.1), the Stringency Index depends mainly on NPIs that decrease contacts with susceptible hosts, and we therefore assume that NPIs only affect transmission, and not the infectious period. We model the link between the effectiveness of the control measures *c*(*t*) and the Stringency Index *Ψ*(*t*) at each time point *t* through the following function:

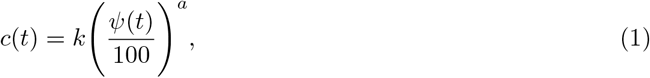

with *k*, the maximum achievable effectiveness, and *a*, a “*shape*” parameter; *Ψ*(*t*) takes values between 0 (no control) and 100. We generated daily new fatality cases (4), daily new cases tested negative (6) and daily new cases tested positive (7) that we fitted to observed data using weighted least squares (WLS) (see **Methods, §4.2.2**). The best WLS estimates for this model yielded *k* = 1 and *a* = 3.78. The adjusted model seemed to fit the general dynamics of the data even though somewhat locally perfectible (**Fig. S6**). We then quantified the uncertainty of our parameter estimates using wild bootstrap [29, 24]: we reiterated about 2000 non-linear optimizations on perturbed data in order to get 2000 new sets of estimations (cf. **Methods, §4.2.2**). We thus obtained the joint distributions of the estimated parameters (see **Fig. S4**), and in particular parameters *k* and *a* that govern equation (1).

In the second step of the analysis we seek to explain the rapid spread of the Alpha variant through an increase in the transmission rate and/or the recovery rate. We developed an *SEIR* model which takes into account the circulation of both the Alpha variant and the previous lineage, which we will refer to as the resident strain (**Methods, §4.1.2**). This model was used to derive an approximation of the temporal dynamics of the overall frequency 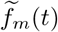 of the Alpha variant. Under the assumptions of weak selection and quasi-equilibrium of fast variables (for more details, see **SI Appendix**), we obtained the following approximation for the selection coefficient s(t) of the Alpha variant:

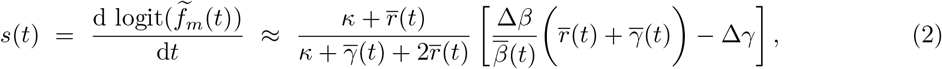

with logit 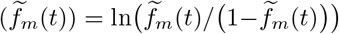 and where Δ*β* and Δ*γ* are the phenotypic differences between the Alpha variant and the resident strain in terms of transmission and recovery, respectively; 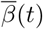 and 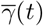 refer to the average transmission and recovery rates across all genotypes; *κ* is the transition rate from the exposed state *E* to the infectious state *I*. Lastly, 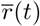 is the average growth rate of the epidemic:

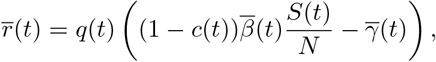

with *q*(*t*), the frequency of infectious individuals among infected hosts (i.e. *I*(*t*)*/ E*(*t*) + *I*(*t*)). It is important to note that NPIs affect the selection coefficient of the variant (2) through the growth rate of the epidemic 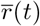, which depends on the amount of control *c*(*t*). Crucially, this impact is stronger if the Alpha variant is more transmissible (i.e. Δ*β >* 0) (see also [9] and [32]). Interestingly, we found – as in [32] – a negative correlation between the selection coefficient of the Alpha variant in England and the Stringency Index: -0.88 at the national scale (95% CI [-0.98; -0.39]) and between -0.97 (London, 95% CI [-0.99; -0.86]) and -0.81 (South West, 95% CI [-0.97; -0.14]) at the regional level (**Fig. S2**). In the following, we approximated 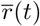 using the quasi-equilibrium expression of *q*(*t*), we assumed that the proportion of susceptible hosts remained approximately constant during the second phase of the analysis (*S*(*t*)*/N ≈ S/N*) and we neglected the effect of the rise in frequency of the variant on the average phenotypic trait values in (2) and 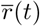 (weak selection assumption).

Under these assumptions along with the previous best WLS estimates for the control parameters from the first step of the analysis (*k* = 1, *a* = 3.78), a linear mixed-effects model (MEM) led to the following estimations of the phenotypic differences (per day): Δ*β* = 0.15 (95% CI [0.033; 0.258]) and Δ*γ* = *−*0.047 (95% CI [*−*0.099; +0.001]) (**Fig. 2**). With a significance level of 5%, likelihood-based comparisons of nested MEMs show a significant effect for Δ*β* but not for Δ*γ* (although with a p-value very close to the significance threshold) (**Table S4**). In addition, we sought to propagate to the second phase the uncertainty of our estimates of the parameters *k* and *a*. Starting from each of the almost 2000 pairs {*k*; *a*} based on previous wild bootstrap computations, we obtained as many new estimators for {Δ*β*; Δ*γ*}. For Δ*β*, 95% of them were between 0.147 and 0.153 (**Fig. S7-A**), for which each corresponding 95% CI remained positive (**Fig. S8**). In contrast, 95% of these 2000 estimates were between -0.054 and -0.046 for Δ*γ* (**Fig. S7-B**), among which 61% of the corresponding 95% CIs included 0 (**Fig. S8**). These distributions led us to conclude that the Alpha variant has a higher transmission rate than the resident strain. With these estimates of Δ*β* and Δ*γ* and in the absence of NPI, the selection coefficient of the Alpha variant was computed, on average, around 0.77 per week (standard deviation: 0.02 per week).

**Figure 2:**
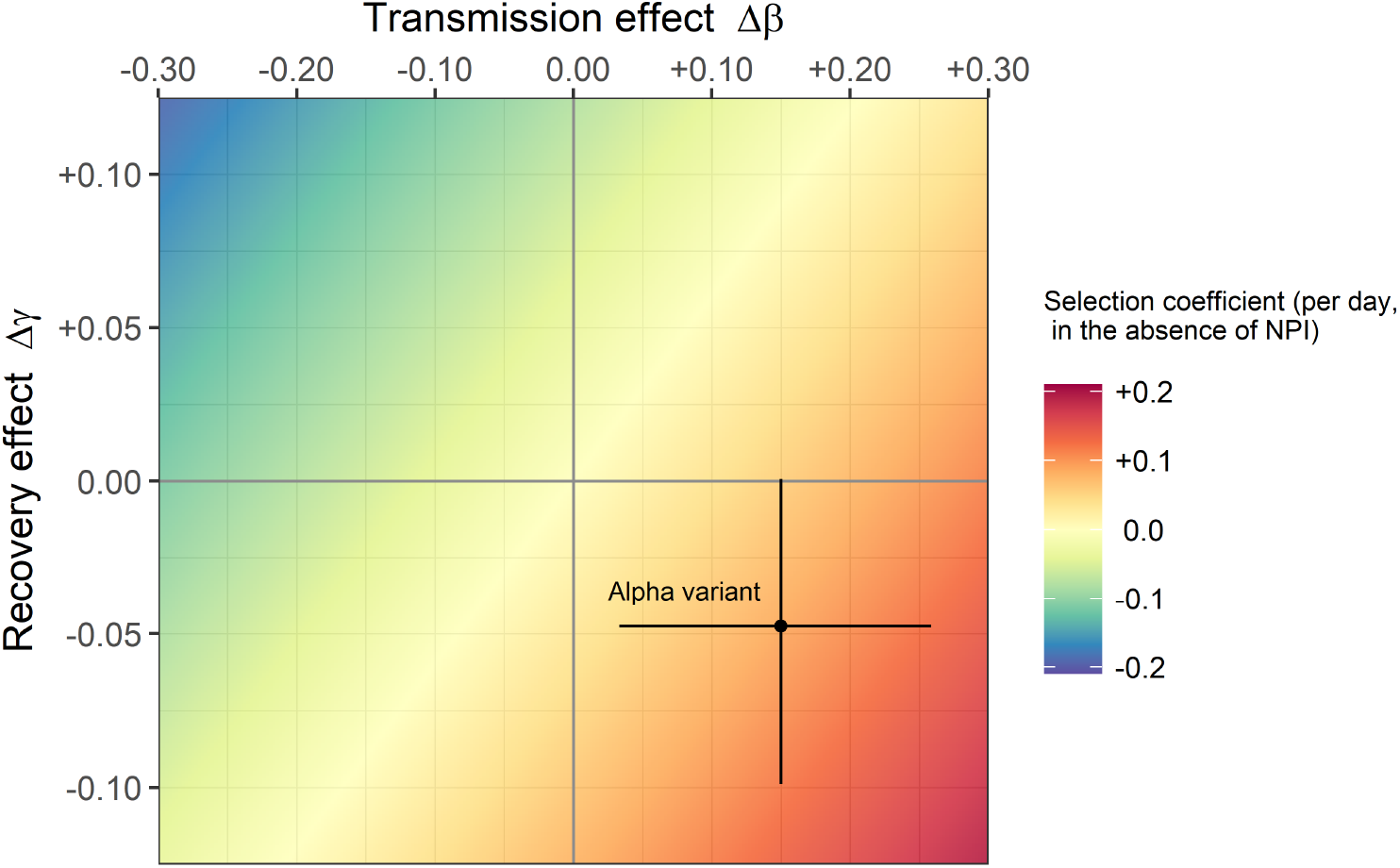
Phenotypic profile of the Alpha variant (transmission and recovery rates) relative to the resident strain. By definition, the phenotype of the resident strain is located at the origin of the graph (Δ*β* = 0; Δ*γ* = 0). Linear MEM estimates (black point, expressed per day) of phenotypic differences in transmission Δ*β* and in recovery Δ*γ* as well as 95% CIs (black cross) are based on the best WLS estimates of control parameters *k* and *a* from the analysis of phase 1 (*k* = 1 and *a* = 3.78). We obtained Δ*β* = 0.15 (95% CI [0.033; 0.258]) and Δ*γ* = *−*0.047 (95% CI [*−*0.099; +0.001]). For the fixed parameters, we set: *S/N* = 0.75, *κ* = 0.2, *β*_*w*_ = 0.25 and *γ*_*w*_ = 0.1. The colored background represents the values of the selection coefficient (in the absence of NPI) as a function of Δ*β* and Δ*γ*; the selection coefficient is here around +0.11 per day (or +0.77 per week) for the Alpha variant. Estimates and 95% CIs based on the joint distributions of parameters *k* and *a* from wild bootstrap computations are represented in **Fig. S8**.

We also explored the robustness of these estimations by applying *±*10% and *±*20% perturbations in the fixed parameters of our model (cf. **Table 1**) to investigate how they would affect our results. First, we kept the best WLS estimates for the control parameters (*k* = 1, *a* = 3.78), and we applied the perturbations to the fixed parameters of the second phase of the analysis. Our estimations of Δ*β* and Δ*γ* were not very sensitive to these perturbations (cf. **Fig. S10**). Second, we applied the perturbations in the fixed parameters of the first step in order to get new estimates of the control parameters *k* and *a*. We used these new estimates in the second phase of the analysis to estimate the phenotypic differences Δ*β* and Δ*γ*. The parameter *k* was hardly affected by these perturbations but the parameter *a* was more sensitive, in particular when varying the transmission rate or the initial proportion of susceptible hosts (cf. **Fig. S11-1**). Next, we reiterated the second step with these new estimations of the pair {*k*; *a*}. All the 95% CIs of the estimates of Δ*β* remained positive after these perturbations. However, some perturbations led to more negative values of Δ*γ* (i.e. the 95% CIs of Δ*γ* included only negative values, **Fig. S11-2**). Note that this effect seems to be driven by the variations in the estimation of the parameter *a* (cf. **Fig. S11**). Taken together, the results of these analyses confirm the conclusion that the Alpha variant has a higher transmission (Δ*β >* 0). An increase in the mean duration of infectiousness (Δ*γ <* 0) seems less likely but cannot be completely ruled out.

**Table 1:**
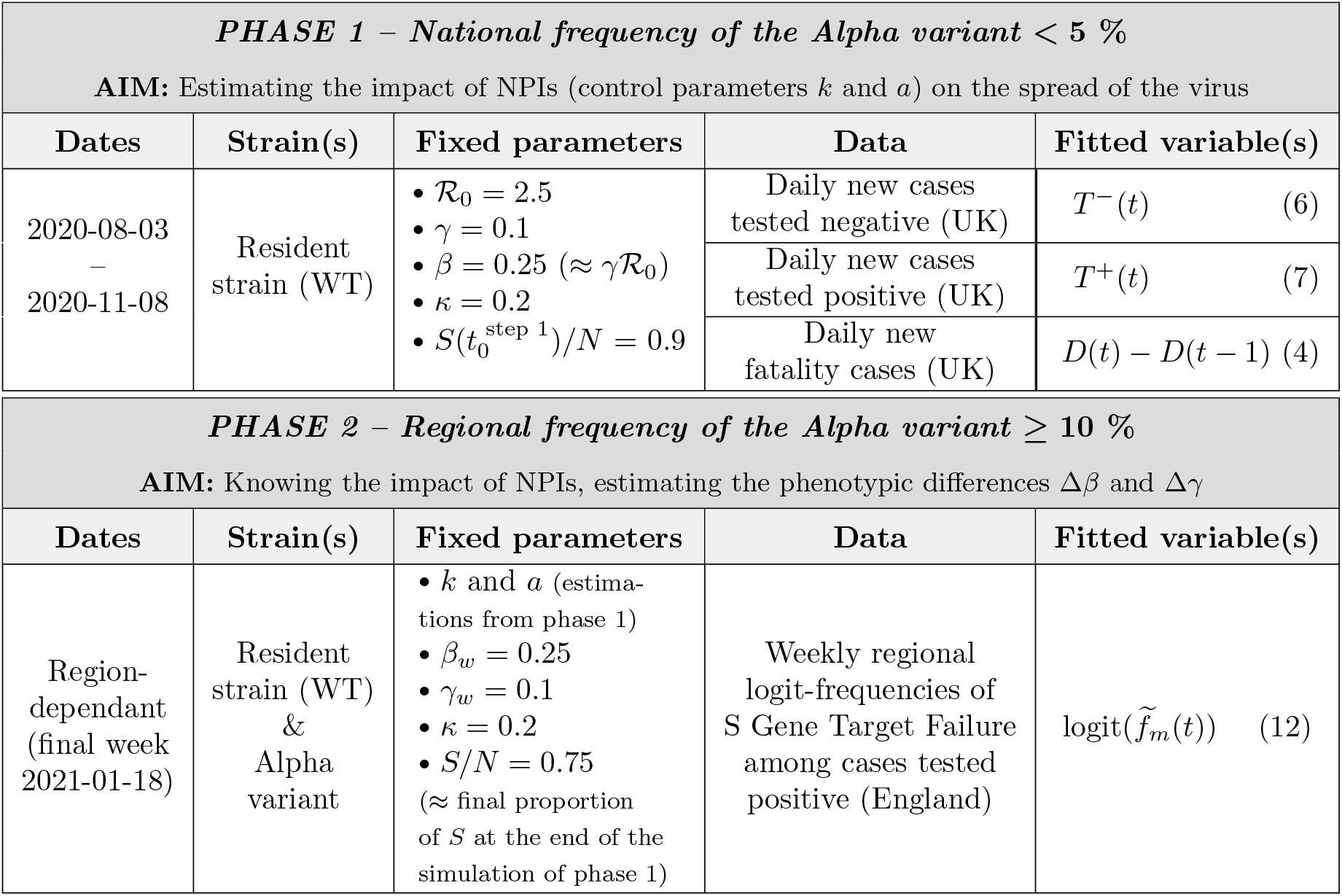
Overview of the two-step analysis. This table summarises the main features of the two phases of the analysis. For each one, we recall the aim, dates, circulating SARS-CoV-2 strains that we considered, fixed parameters, data and fitted variables (model) – equation numbers are specified between brackets just after the corresponding variable. For both phases, we also use values of the Stringency Index in the UK. ℛ_0_, *γ* (or *γ*_*w*_) and *β* (or *β*_*w*_) are the basic reproduction number and the *per capita* rates (per day) of recovery and transmission, respectively, of the resident strain *w*; *κ* is the *per capita* transition rate (per day) from the exposed to the infectious state (same for both strains); *k* and *a* are the parameters linking the Stringency Index to the efficacy of NPIs (same for both strains); *S*(*t*)*/N* is the proportion of susceptible hosts in the population (assumed constant in the second phase); *D* refers to the cumulative density of COVID-19-related deaths. See **Table S1** and **S3** for a more detailed summary of the parameters involved in phase 1 and 2, respectively.

| PHASE 1 – National frequency of the Alpha variant < 5 % |  |  |  |  |
| --- | --- | --- | --- | --- |
| AIM: Estimating the impact of NPIs (control parameters $k$ and $a$) on the spread of the virus | | | | |
| Dates | Strain(s) | Fixed parameters | Data | Fitted variable(s) |
| 2020-08-03<br>–<br>2020-11-08 | Resident strain (WT) | • $\mathcal{R}_0 = 2.5$<br>• $\gamma = 0.1$<br>• $\beta = 0.25$ ($\approx \gamma \mathcal{R}_0$)<br>• $\kappa = 0.2$<br>• $S(t_0^{\text{step } 1})/N = 0.9$ | Daily new cases tested negative (UK) | $T^-(t)$ (6) |
| | | | Daily new cases tested positive (UK) | $T^+(t)$ (7) |
| | | | Daily new fatality cases (UK) | $D(t) - D(t - 1)$ (4) |
| PHASE 2 – Regional frequency of the Alpha variant $\geq 10$ % | | | | |
| AIM: Knowing the impact of NPIs, estimating the phenotypic differences $\Delta\beta$ and $\Delta\gamma$ | | | | |
| Dates | Strain(s) | Fixed parameters | Data | Fitted variable(s) |
| Region-dependant (final week 2021-01-18) | Resident strain (WT) & Alpha variant | • $k$ and $a$ (estimations from phase 1)<br>• $\beta_w = 0.25$<br>• $\gamma_w = 0.1$<br>• $\kappa = 0.2$<br>• $S/N = 0.75$ (≈ final proportion of $S$ at the end of the simulation of phase 1) | Weekly regional logit-frequencies of S Gene Target Failure among cases tested positive (England) | $\text{logit}(\tilde{f}_m(t))$ (12) |

## 3 Discussion

We developed a two-step approach to characterise the phenotypic variation of the Alpha variant relative to the previously dominant lineage. In the first step of the analysis, we focus on the epidemiological dynamics before the emergence of the Alpha variant and we used an *SEIR* model, a simplified representation of an age-structured model, to infer the effect of the Stringency Index on the reduction of transmission induced by these control measures. This led us to infer a convex increasing function that captures the effect of the Stringency Index on the reduction in the number of contacts with susceptible hosts (**Fig. S5**).

The second step of this approach is based on the analysis of the change in frequency of the Alpha variant after its emergence. Using evolutionary epidemiology theory [8, 7, 9], we derive an expression for the gradient of selection in an *SEIR* model. The analysis of selection in such a class structured environment (the virus is infecting both the *E* and the *I* hosts) is facilitated under the assumption of weak selection and the approximation of quasi-equilibrium for fast variables [27, 14, 28]. We recover a classical result derived in simpler *SIR* models: the intensity of selection for higher transmission rates depends on the availability of susceptible hosts and the amount of NPIs aiming to reduce contact (e.g. social distancing or face coverings). In contrast, selection for longer durations of infectiousness is much less sensitive to these control measures. Using our independent estimation of the effectiveness of NPIs based on the Stringency Index, we inferred both Δ*β* and Δ*γ* of the Alpha variant from the temporal dynamics of its logit-frequency. This analysis suggests that the selective advantage of the Alpha variant was mainly driven by a higher transmission rate. An increase in the mean duration of infectiousness (i.e. a lower rate of recovery) seems less likely but cannot be completely ruled out. Interestingly, recent experimental studies of viral transmission confirm the transmission advantage of the Alpha variant. Viral shedding in breath aerosols were recently found to be higher in individuals infected with the Alpha variant than with previous lineages [25].

Several specific mutations of the Alpha variant could explain these phenotypic differences. Preliminary genomic characterisations detected around 17 non-synonymous substitutions or deletions compared to the previous lineage; about half were associated with the protein S gene, including mutations of immunological significance [38]. In particular, the mutation N501Y, known to increase the affinity of the viral glycoprotein S for the human receptor ACE2 [44], and the mutation P681H, adjacent to a serine protease cleavage site that is required for cell infection [23], are both likely to affect the withinhost development of the virus in infected hosts. How this development affects key phenotypic traits like transmission and recovery rates in human host is difficult to explore experimentally. Our analysis can thus provide a complementary approach that may help to link genetic and phenotypic variation.

Yet, it is important to note that this analysis relies on several simplifying assumptions. For instance, we assumed that infectiousness began at the same time as the onset of symptoms – i.e. the latent period and the incubation period coincide perfectly in time. Yet, transmission from a pre-symptomatic state is a distinctive feature of SARS-CoV-2 [41, 9, 20]. Besides, our framework sticks to the *SEIR* class of models formalised by ODEs, with *κ* and *γ*, the (constant) rates of leaving the exposed and infectious compartments, respectively. This implicitly yields sojourn times in the different compartments that are exponentially distributed – and thus, markovian or memoryless [13, 43]. As a result, the generation time follows a hypoexponential distribution (generalized Erlang distribution) with mean 1*/κ* +1*/γ* and variance 1*/κ*^2^ + 1*/γ*^2^ [48]. Our analysis does not allow the mean and variance of this distribution to change independently but a variation in *γ* does affect the mean and the variance of the generation time. Several studies, however, have discussed the influence of the shape of the generation time distribution on both the epidemiological and evolutionary dynamics of the pathogen [6, 48, 34, 33, 3, 1]. We show in the **SI Appendix §S7** how to recover our results using the selection on the shape of the generation time distribution used by Blanquart *et al*. [3]. In both analyses, variations in the intensity of NPIs are assumed to impact the effective reproduction number without altering the generation time distribution (which means they only impact transmission). Nevertheless, some control measures like contact tracing and post-symptomatic isolation may impact the duration of infectiousness, the generation time distribution and the selection on the variant [33].

Data availability and quality are major limiting factors in any statistical inference analysis. The Stringency Index provides a rough approximation of the intensity of control at the national scale. More precise and more local estimations of control would allow us to refine our estimations. In addition, we show in the **SI Appendix §S6** how the availability of data frequency among different types of hosts (i.e. the differentiation between the exposed and the infectious compartments) may provide another way to estimate Δ*β* and Δ*γ*.

To conclude, we contend that it is important to exploit the joint epidemiological and evolutionary dynamics of SARS-CoV-2 to better understand its phenotypic evolution. This phenotypic evolution is undermining our efforts to control the epidemic. New variants are emerging and are affecting other phenotypic traits. In particular, the ability of new variants (e.g. Omicron) to escape immunity has major impact on the epidemiological dynamics [35]. Inference approaches using both epidemiological and evolutionary analysis could yield important insights on the adaptive trajectories on the phenotypic landscape of SARS-CoV-2, and possibly other pathogens.

## 4 Methods

### 4.1 A two-step analysis

The analysis is performed in two steps considering two consecutive evo-epidemiological periods of time: before and after the emergence of the Alpha variant in England (**Fig. 1**). The first step aims to estimate the force of infection in the presence of NPIs. In particular, we quantify *c*(*t*), a function measuring the impact of NPIs at time *t* on the force of infection *λ*(*t*). This first step takes place temporally before the emergence of the Alpha variant – i.e. before it reaches 5% of the cases tested positive in England – and consists in modeling the epidemiological phase of the previous lineage, which we refer to as the resident strain, disregarding the pre-existing genetic diversity [22]. The second step consists in estimating the differences in contagiousness and in infectious duration in the presence of NPIs during the period when the two strains cohabit – i.e. for each region, from the moment the frequency of the variant reaches 10% of cases tested positive. We combine information from screening and mortality data for the first step (using an epidemiological model), while we focus on the changes in frequency of the variant among positive cases for the second step. See **Table 1** for an overview of this two-step approach.

For both steps, we consider a host population of size *N*. We note *S, E, I* and *R*, respectively, the states (or compartments) of individuals that are <u>S</u>usceptible to the disease, <u>E</u>xposed – i.e. infected but not yet infectious -, <u>I</u>nfectious and <u>R</u>ecovered. For a given state, for instance *S*, and current time *t* (expressed in days), we note *S*(*t*) the density of people in that state and 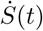 its differentiation with respect to time. Let *β* be the *per capita* transmission rate (direct and horizontal) and *γ* the *per capita* recovery rate. Control measures implemented by governments such as social distancing, face coverings, lockdowns or travel bans are NPIs that aim to curb the spread of the epidemic by alleviating the force of infection *λ*(*t*) = *βI*(*t*)*/N*. Given *c*(*t*) the effectiveness of these measures – ranging from 0 (no control) to 1 (total control) –, the expression for the force of infection thus becomes: *λ*(*t*) = (1 *− c*(*t*))*βI*(*t*)*/N*. Directly estimating the control efficiency *c*(*t*) is usually impossible; it results from a multitude of factors that may vary spatially and temporally and is not necessarily proportional to the severity of the measures in place. This is why we choose here to include the *Stringency Index* (which we noted *Ψ*(*t*)), a composite score published by OxCGRT [18]. This index is based on nine component indicators and rescaled to a value between 0 (no control) and 100 (the stringest) in order to reflect the strictness of public health policy. Eight component indicators are related to “*containment and closure*” (school and workplace closing, cancel public events, restrictions on gathering site, close public transport, stay-at-home requirements and restrictions on internal movement and on international travel) and one is related to “*health system*” (public information campaign) [18]. These measures, in contrast with post-symptomatic isolation or contact tracing (not explicitly taken into account in this score), are mainly limiting the number of contacts *un*conditionally to infection, that is mostly intended to reduce the transmission rate than to shorten the infectious period. We thus assume that NPIs included in the Stringency Index would only affect the transmission rate (and not the infectious period). Although somewhat imperfect, this index has the advantage of integrating many factors into one value, as well as being available per day online since the onset of the pandemic in many countries. We model the link between *c*(*t*) and *Ψ*(*t*) through the following concave or convex relationship:

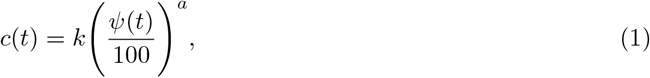

with *k ∈* [0; 1], the maximum achievable efficiency (when *Ψ*(*t*) = 100), and with *a* 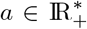, a ‘*shape*’ parameter.

#### 4.1.1 Step 1: epidemiological analysis just before the emergence of the Alpha variant

We use a version of the well-known *SEIR* model (see **Fig. S3**) to estimate the parameters that govern the epidemiological dynamics before the arrival of the Alpha variant. We denote *α*, the additional *per capita* mortality rate induced by the viral disease (i.e. the virulence) and *D*, the compartment of (COVID-19-related) deceased individuals. We assume that the (potential) onset of symptoms and the onset of infectiousness occur simultaneously after a latent period of mean duration 1*/κ*. Within the infectious compartment *I*, some hosts develop symptoms (*I*_*S*_) with probability *ω* while the others remain asymptomatic (*I*_*A*_) with complementary probability. It is further assumed that individuals *I*_*A*_ systematically recover at a *per capita* rate *γ* while individuals *I*_*S*_ are divided into two sub-compartments depending on their fate: *I*_*Sd*_, with probability *p*, for those who will eventually die from the disease (with virulence *α*), or, alternatively, *I*_*Sr*_, for those who will eventually recover (at the same rate *γ* as asymptomatic hosts). We model these epidemiological trajectories using the following system of ODEs:

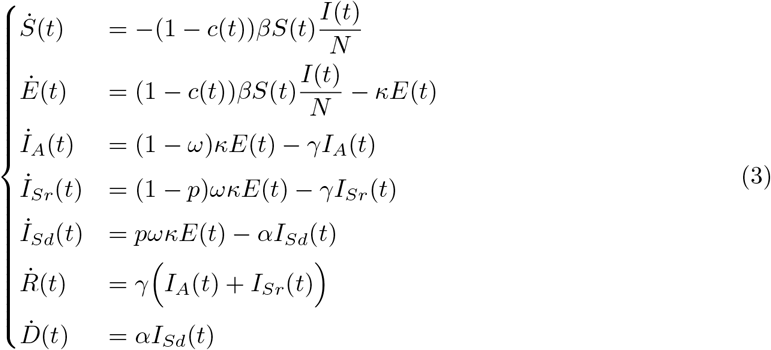

Following [10] for the construction of the Next Generation Matrix, the basic reproduction number ℛ_0_ – i.e. the expected number of *infectees* from one *infector* in a fully susceptible population – is then given in the absence of NPI by:

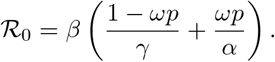

In the context of COVID-19, the product *ωp* – i.e. the probability of developing symptoms and dying from the disease – is very low. We then approximate the basic reproduction number of the resident strain of SARS-CoV-2 as ℛ_0_ *≈ β/γ*.

At each time point (each day), only a small fraction of the population is tested and hosts with symptoms are more likely to be tested than others. In order to take these biases into account, we use the following range of assumptions:

- Individuals *S* and *I*_*A*_ are tested with the same probability / reporting rate *ρ*;
- Individuals *S* and *I*_*A*_ can be tested several times;
- All new individuals *I*_*S*_ (symptomatic) are tested (reporting rate of 1);
- Screening of individuals *E* and *R* is neglected (reporting rate of 0);
- All new disease-related deaths are reported (reporting rate of 1).

Furthermore, screening efforts in the UK tended to be strengthened over time during this period (as shown for instance by the increasing number of negative tests in **Fig. S6**). As the reporting rate for individuals without symptoms *S* and *I*_*A*_ can no longer be considered constant, we also assume a linear increase with time:

- The reporting rate *ρ* for individuals *S* and *I*_*A*_ (without symptoms) increases linearly over time: *ρ*(*t*) = *η t* + *μ*.

The reporting rate is not identifiable in an *SIR* model when only a fraction of the compartment *I* is observed [19]. Thus, we also consider the disease-related deaths in the observation process. The combination of information, that is daily new cases tested negative and tested positive and daily new fatality cases, allow us to identify the reporting rate. Between two consecutive time points *t −* 1 and *t*, the number of new fatality cases is given by:

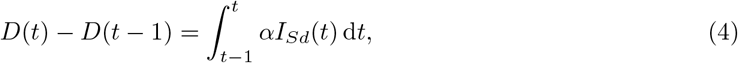

and, given 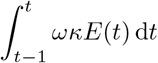, the *incidence* of symptomatic cases (i.e. new incomers in compartment *I*_*S*_), we decomposed the number of performed tests *T* (*t*) as follows:

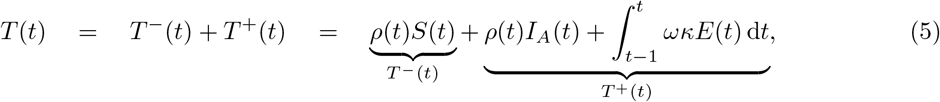

with *T* ^*−*^(*t*) and *T* ^+^(*t*), the number of cases tested negative and tested positive, respectively. Thus:

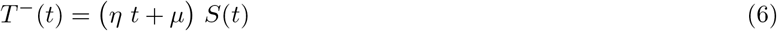

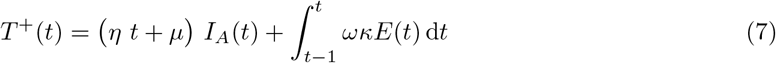

#### 4.1.2 Step 2: Evolutionary analysis

We now consider that two distinguishable pathogenic strains compete: the resident (or WT) strain, represented with the subscript *w*, and the mutant strain (or variant), represented with the subscript *m*. The total number of exposed hosts *E*(*t*), where *t* is the current time, can therefore be decomposed into: *E*(*t*) = *E*_*m*_(*t*) + *E*_*w*_(*t*). Likewise, for the infectious hosts *I*(*t*): *I*(*t*) = *I*_*w*_(*t*) + *I*_*m*_(*t*), and we denote *q*_*m*_(*t*) = *I*_*m*_(*t*)*/I*(*t*), the frequency of the variant in *I*. We propose that the variant may differ phenotypically from the resident strain in its effective transmission rate *β*_*m*_ = *β*_*w*_ + Δ*β* and/or its recovery rate *γ*_*m*_ = *γ*_*w*_ + Δ*γ*. In contrast, we neglect any difference in terms of latent period (*κ*_*m*_ = *κ*_*w*_ = *κ*), and we neglect the virulence of both strains (*α*_*m*_ = *α*_*w*_ = 0). For SARS-CoV-2, frequencies of the Alpha variant did not seem to depend on the age of hosts [5]. Assuming furthermore that over-infections do not occur – including co-infections with both strains – and that (persistent) immunity acquired with either strain protects effectively against both, we start with the simple following *SEIR* model:

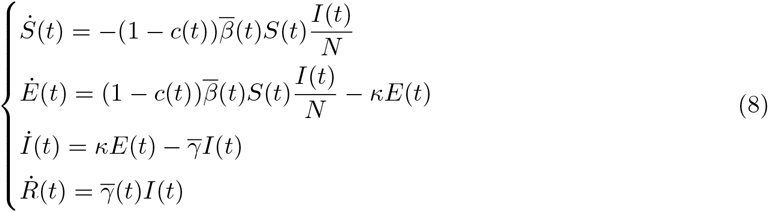

where the overlines refer to mean values of the phenotypic traits after averaging over the distribution of strain frequencies:

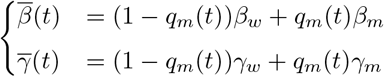

As described in [27, 28], under the assumption of weak selection, the overall frequency of the variant 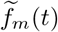 can be tracked using:

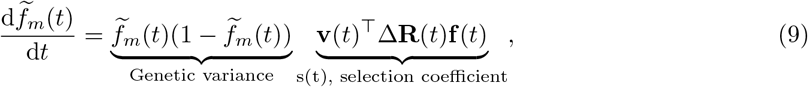

with **v**(*t*) and **f** (*t*), the vectors of reproductive values and class frequencies, respectively, within the infected states (*E* and *I*), and Δ**R**(*t*), the matrix of differences in transition rates between the mutant strain and the resident strain (for more details, see **SI Appendix**). An easier way to study *s*(*t*) in time series analyses is not to directly work with frequencies but with logit-frequencies instead, that is ln(frequency of the variant / frequency of the resident strain). Indeed, it may easily be shown that:

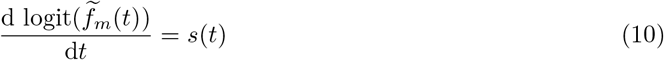

We then focus on the selection coefficient of the variant *s*(*t*) (also known as the selection gradient). According to its value (weakly or strongly positive, weakly or strongly negative), this selection coefficient quantifies over time the success or the disadvantage of the variant over the resident strain through natural selection [8, 7, 9]. In the **SI Appendix**, we show that, using quasi-equilibrium approximations for fast variables, the selection coefficient of the variant *s*(*t*) may be approximated as:

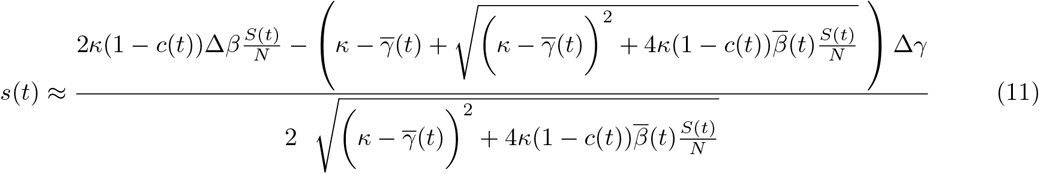

For the *SIR* model nested in the *SEIR* model (8), the selection coefficient is merely: *s*(*t*) = (1 *− c*(*t*))Δ*βS*(*t*)*/N −* Δ*γ* [8, 7], which shows analytically the importance of the control through *c*(*t*) to distinguish the scenario where the selective advantage of the variant stems from a higher transmission rate (Δ*β >* 0; Δ*γ* = 0) from the scenario with a longer duration of infectiousness (Δ*β* = 0; Δ*γ <* 0), or from an intermediate scenario (Δ*β* ≠ 0; Δ*γ* ≠ 0). In other words, it is particularly the variations in *c*(*t*) that might help us to decouple the effects of these two phenotypic traits. Simply adding an exposed state *E* makes the selection gradient surprisingly much more difficult to express but the importance of the variations in *c*(*t*) for this purpose (although less clear-cut) remains nevertheless relevant as suggested by (11).

### 4.2 Statistical inference

#### 4.2.1 Programming

Numerical simulations and data analyses were carried out using R [37] version 4.1.1 (2021-08-10). ODEs were solved numerically by the function ‘*ode*’ (method ‘*ode45*’) from the package ‘*deSolve*’ [42].

#### 4.2.2 Step 1

We used daily screening data between 2020-08-03 and 2020-11-08 in the UK (a period for which the Alpha variant was below 5% among cases tested positive in England); 7-day rolling average data were used in order to mitigate the effects of variation in testing activity, e.g. during weekends. We also included daily COVID-19-related deaths in the UK (‘*Daily deaths with COVID-19 on the death certificate by date of death*’) as well as the Stringency Index.

The goal of this part is to compute *c*(*t*) from the Stringency Index and thus to focus on the estimation of the parameters *k* and *a*. We used additional information from the literature to fix the value of some parameters of the model (3): we set the mean latent period to 5 days [11] and the mean duration of infectiousness to 10 days [4] – that is, an average infection period of 15 days –; we also set the basic reproduction number ℛ_0_ to 2.5 [12, 26] and the initial proportion of susceptible hosts to 0.9. Besides, we approximated the initial states within compartment *I*. This is summarised with further details in **Table S1**. With these parameters fixed, the model (3) is identifiable (**Fig. S12**, following [39]). The remaining parameters of the first phase were estimated using weigthed least squares (WLS). Let 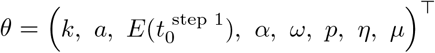 be the vector of parameters to estimate, with 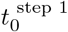 the initial time point of the first step, and 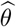 its estimator such that:

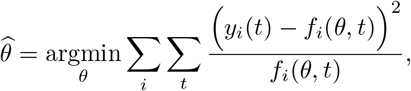

where the subscript *i* refers to our three observation states – i.e. daily new cases tested negative and tested positive and daily new fatality cases –, which were modelled through functions *f*_*i*_. *f*_*i*_(*θ, t*) corresponds thus to the expected observations while *y*_*i*_(*t*) corresponds to the real observations (data). With WLS, squared residuals are weighted by the inverse of the variance of the observations *y*_*i*_(*t*); these weights balanced the contrasting intrinsic contributions of each observation – e.g. negative tests and deaths are not on the same order of magnitude. Assuming *y*_*i*_(*t*) to be Poisson-distributed – consistent with ODEs where sojourn times are exponentially distributed – then the variance of the observations is *f*_*i*_(*θ, t*). This would correspond to the Pearson *χ*^2^ function in [2]. Non-linear optimizations were tackled with the R function ‘*optim*’, from the basic package ‘*stats*’, using the Nelder-Mead (or downhill simplex) method – maximum number of iterations *maxit* = 2000, absolute and relative convergence tolerance *abstol* = *reltol* = 10^*−*6^. This optimization procedure was iterated for 1500 sets of uniformly drawn initial values (because of the presence of local minima) and was restricted to certain ranges of values through parameter transformations (cf. **Table S2**). Only parameter estimates from the best fit – i.e. successful completion with the lowest WLS value – were kept and we refer to them as the best WLS estimates.

Parameter distributions were then computed using wild bootstrap [29, 24], which allow in particular to take into account any heteroscedasticity in the residuals. To do this, 2000 sets of bootstraped data were generated: residuals were perturbed by an i.i.d. sequence of *n* random weights 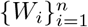 following Mammen’s 2-points distribution (that is, 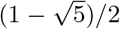 with probability 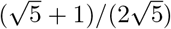 and 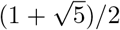 with probability 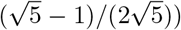, which satisfies 𝔼 (*W*_*i*_) = 0 and 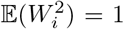 [24]. Non-linear optimizations were then reiterated, but starting only from the best WLS estimates and the corresponding set of initial values.

As a sensitivity analysis, *±*10% and *±*20% perturbations were applied to the fixed parameters of the first phase separately (*β, κ, γ* and 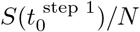) and non-linear optimizations were each time reiterated starting from a set of 500 initial conditions (uniformly drawn, as before).

#### 4.2.3 Step 2

We used weekly regional frequencies of S Gene Target Failure (SGTF) in England from the technical briefing 5 of Public Health England (PHE), which was investigating the new VOC 202012/01 variant between September 2020 and January 2021 [36]. Briefly, qPCR from the ThermoFisher TaqPath kit (designed to target three genes: ORF1ab, N and S) were performed after swab sampling in the the wider population – i.e. outside NHS hospitals and PHE labs. Due to the deletion ΔH69/V70 in the genome of the Alpha variant, a mismatch between one of the three molecular probes and the viral sequence encoding for the glycoprotein Spike (S) resulted in a failure of detection, or SGTF, a genomic signature that was then used as a proxy for this variant [36, 47]. As in the first step, we also included values of the Stringency Index in the UK.

Under the assumption that variations in *S*(*t*)*/N* on short time scales may be neglected for a controlled epidemic (*S*(*t*)*/N ≈ S/N*) and by neglecting the effect of the rise in frequency of the variant on the average phenotypic trait values – i.e. 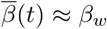 and 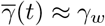 (weak selection approximation) –, we may integrate (11) in accordance with (10) to find an expression for the overall logit-frequency of the variant:

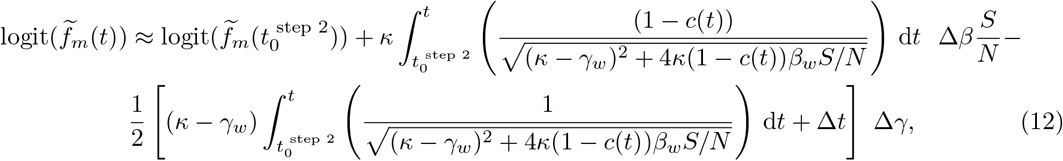

where 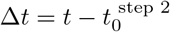 is the period of time between the system at time *t* and its initial state.

Δ*β* appears as a product with *S/N* in (12), which implies that they are likely not to be separately identifiable. At the final time point of the first phase, our best fit ended up with a proportion of susceptible hosts around 0.75. Hence, we consistently set *S/N* to 0.75 for the second phase. As in the first phase, we also set: *κ* = 0.2, *β*_*w*_ = 0.25 and *γ*_*w*_ = 0.1. Phenotypic differences relative to the previous lineage (Δ*β* and Δ*γ* in (12)) were estimated using a linear mixed-effects model (MEM) to fit weekly logit-frequencies of SGTF among cases tested positive to COVID-19 as a proxy of the Alpha variant in the nine regions of England late 2020 early 2021. We assumed that these frequencies were representative of the infected population and that the regions were independent of each other – i.e. no inter-region flows. In more detail, 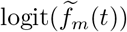 was the response variable, Δ*β* and Δ*γ* were treated as fixed effects and the region (nine in total) was treated as a random effect on the intercept of the model. Hence, for the region *i* at time point *t* (*i* and *t* are now noted as indexes for clarity):

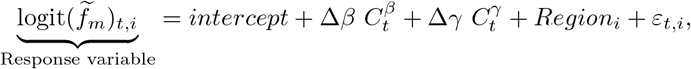

with:

- *intercept*, the common fixed effect (reference);
- 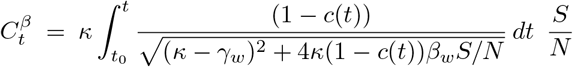, the covariate associated with Δ*β* (fixed effect);
- 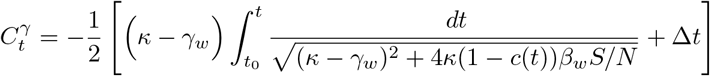, the covariate associated with Δ*γ* (fixed effect);
- *Region*_*i*_ *∼* 𝒩 (0, *ν*^2^), the random effect (with variance *ν*^2^) of the region *i* on the intercept of the model;
- *ε*_*t,i*_ *∼* 𝒩 (0, *σ*^2^), the residual error (with variance *σ*^2^).

This MEM was implemented using the function ‘*lmer* ‘ from the R package ‘*lme4* ‘: logit 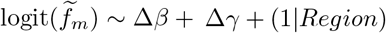, and 95% CIs of parameters Δ*β* and Δ*γ* were computed using the function ‘*confint* ‘ from the package ‘*stats*’. For each region, the initial date corresponds to the moment the Alpha variant reached 10% of cases tested positive – i.e. above horizontal lines in **Fig. S1-D**. Below this threshold, the dynamics of the variant could not be considered as deterministic. The parameters *k* and *a* that govern the link between the Stringency Index the and intensity of control (1) were set according to their best WLS estimates and joint distribution that were previously computed in the first step (cf. **§4.2.2**).

As for the first step, we investigated the robustness of our estimations. First, keeping our best WLS estimates for parameters *k* and *a*, linear MEM were reiterated with *±*10% and *±*20% perturbations in the fixed parameters of the second phase separately (*β*_*w*_, *κ, γ*_*w*_ and *S/N*). Secondly, we used estimations of parameters *k* and *a* that we obtained after perturbing the fixed parameters of the first phase (cf. **§4.2.2**) to propagate these perturbations to the outcomes of the second step; the value of the fixed parameters of the second phase were each time updated in accordance.

## Data Availability

Frequencies data were taken from the Public Health England (PHE) Technical Briefing 5, which was investigating the new VOC 202012/01 variant (Alpha) between September 2020 and January 2021: https://assets.publishing.service.gov.uk/government/uploads/system/uploads/attachment_data/file/957631/Variant_of_Concern_VOC_202012_01_Technical_Briefing_5_Data_England.ods;
daily epidemiological data from national screening in UK were downloaded from the website 'Our World in Data': https://ourworldindata.org/grapher/daily-tests-and-daily-new-confirmed-covid-cases?country=~GBR;
daily COVID-19-related deaths in UK were downloaded from the governmental website 'GOV.UK': https://coronavirus.data.gov.uk/details/deaths (see 'Daily deaths with COVID-19 on the death certificate by date of death');
eventually, the Stringency Index is computed by the Oxford COVID-19 Government Response Tracker: https://www.bsg.ox.ac.uk/research/research-projects/covid-19-government-response-tracker.

https://coronavirus.data.gov.uk/details/death

https://www.bsg.ox.ac.uk/research/research-projects/covid-19-government-response-tracker

## Abbreviations (alphabetical order)

ACE2: Angiotensin-Converting Enzyme 2
CI: Confidence Interval
COVID-19: COronaVIrus Disease-2019
i.i.d.: independent and identically distributed
MEM: Mixed-Effects Model
NHS: National Health Service *(UK)*
NPI: Non-Pharmaceutical Intervention
ODE: Ordinary Differential Equation
ORF1ab: Open Reading Frames 1a and 1b
OxCGRT: Oxford COVID-19 Government Response Tracker
PHE: Public Health England *(now replaced by UKHSA)*
qPCR: quantitative Polymerase Chain Reaction
S: Spike *(viral gene and protein)*
SARS-CoV-2: Severe Acute Respiratory Syndrome-CoronaVirus-2
*S*(*E*)*IR*: Susceptible-(Exposed-)Infectious-Recovered
SGTF: S Gene Target Failure
VOC: Variant Of Concern
WHO: World Health Organization
WLS: Weighted Least Squares
WT: Wild Type

## Author contributions

Conceptualization, Methodology, Investigation and Writing (original draft and review & editing): WB, SL, RC and SG; Formal analysis: WB; Vizualisation: WB and SG; Supervision: SL, RC and SG.

## Data accessibility statement

Data that were used in this study are available in a GitHub repository, along with the scripts for the analyses (in R).

## Acknowledgements

We thank Troy Day and François Blanquart for inspiring discussions.

