## Supplementary Figures and Tables for "Phenotypic evolution of SARS-CoV-2: a statistical inference approach"

March 27, 2023

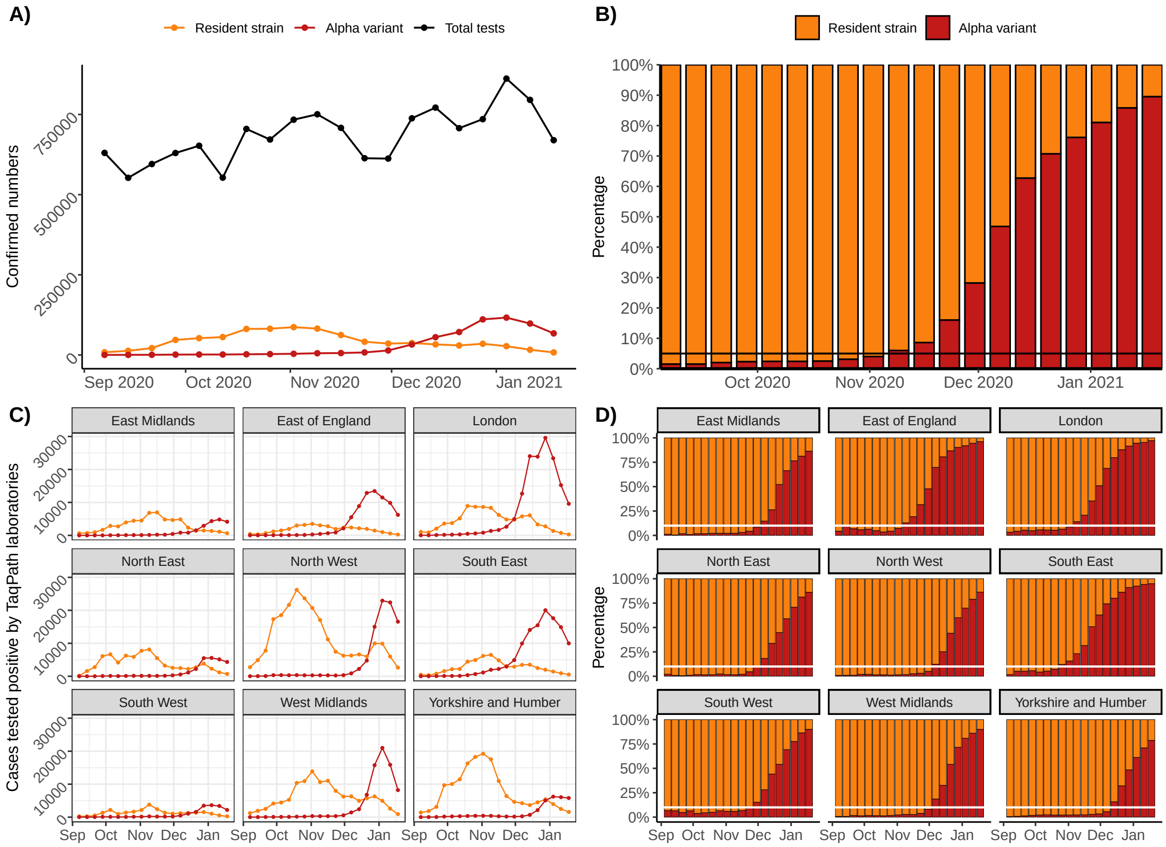


Figure S1: **Epidemiological and genetic data from the COVID-19 outbreak in England between September 2020 and January 2021.** Weekly numbers of TaqPath Pillar 2 COVID-19 positive tests associated with the resident strain of SARS-CoV-2 or with the Alpha variant (lineage B.1.1.7) at the national scale (**A**, total numbers of tests (not only TaqPath tests) are shown in black) and at regional scale (**C**). Weekly percentages of each strain within the TaqPath Pillar 2 COVID-19 positive tests at the national scale (**B**) and at the regional scale (**D**). SGTF from qPCR was used as a proxy of the Alpha variant. In this study, we considered two consecutive evo-epidemiological phases: (i) the phase that preceded the emergence of the Alpha variant, and (ii) the phase that followed it. The first phase took place just before the frequency of the variant reached 5% of the cases tested positive at the national scale (horizontal black line in **B**). Then, for each region, the second phase started at the date the variant reached at least the threshold value 10%, indicated in **D** with horizontal white lines. Below, the number of cases associated with the Alpha variant was quite low and the dynamics of its frequency was widely driven by stochastic processes – see for example the erratic dynamics below the horizontal white line for the second region ('East of England') in **D**.

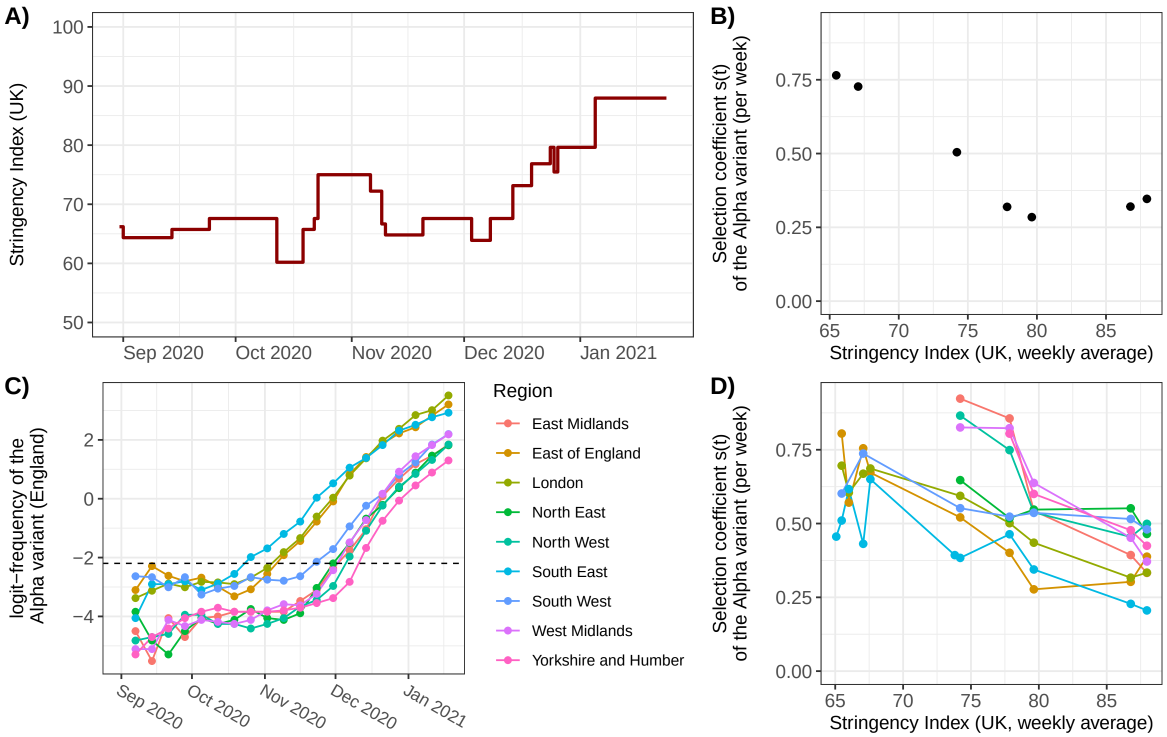


Figure S2: **The selection coefficient of the Alpha variant in England is negatively correlated with the Stringency Index in the UK (fall - winter 2020-2021).** (**A**) Daily values of the Stringency Index in the UK; (**C**) Temporal dynamics of the logit-frequency of SGTF, used as a proxy of the Alpha variant, for each region of England (the black horizontal dashed line indicates the threshold frequency 10%); (**B-D**) Selection coefficient $s(t)$ (per week) of the Alpha variant – i.e. slope of its logit-frequency over time - at the national scale (**B**) and by region (**D**) against the Stringency Index (weekly average) – only frequencies greater than or equal to 10% (above the threshold in **C**) were considered. The correlation between $s(t)$ and the Stringency Index at the national scale is -0.884 (95% CI [-0.983; -0.390]) with a significance test yielding a *p-value* of $8.33 \times 10^{-3}$. Correlations at the regional scale are: East Midlands: -0.948 (95% CI [-0.997; -0.400], *p-value* $= 0.0142$), East of England: -0.868 (95% CI [-0.972; -0.483], *p-value* $= 2.39 \times 10^{-3}$), London: -0.969 (95% CI [-0.994; -0.857], *p-value* $= 1.61 \times 10^{-5}$), North East: -0.885 (95% CI [-0.987; -0.263], *p-value* $= 0.0189$), North West: -0.899 (95% CI [-0.993; -0.080], *p-value* $= 0.0381$), South East: -0.846 (95% CI [-0.959; -0.499], *p-value* $= 1.04 \times 10^{-3}$), South West: -0.809 (95% CI [-0.971; -0.142], *p-value* $= 0.0276$), West Midlands: -0.968 (95% CI [-0.998; -0.588], *p-value* $= 6.8 \times 10^{-3}$), Yorkshire and Humber: -0.931 (95% CI [-0.999; 0.289], *p-value* $=0.0694$).

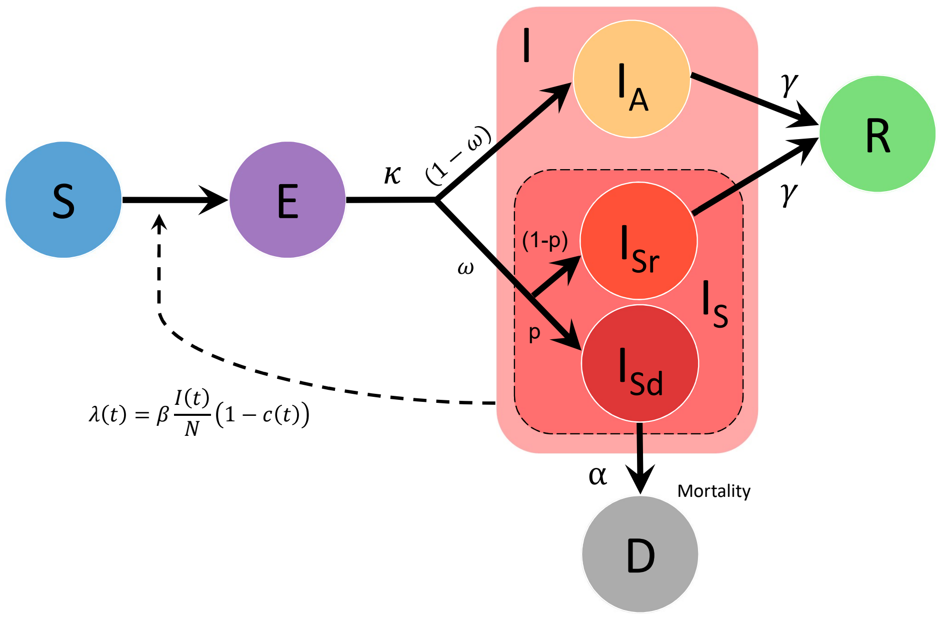


Figure S3: **Flow chart of the epidemiological *SEIR* model used in the first step of our analysis (phase 1)**. Hosts may be: $S$ (Susceptible to the infection), $E$ (Exposed, that is infected but not yet infectious), $I$ (Infected, with $I_A$: Asymptomatic; $I_S$: Symptomatic (with subscript $r$ for those for will eventually recover and $d$ for those who will eventually die from the disease)), $R$ (Recovered and immunised) and $D$ (Deceased). Transitions are represented by solid line arrows associated with the corresponding parameters (see Table S1 for definitions). The dashed line arrow symbolises the role of compartment $I$ in the force of infection $\lambda(t)$ – i.e. transition rate from compartment $S$ to compartment $E$. $c(t)$ is the efficacy of NPIs.

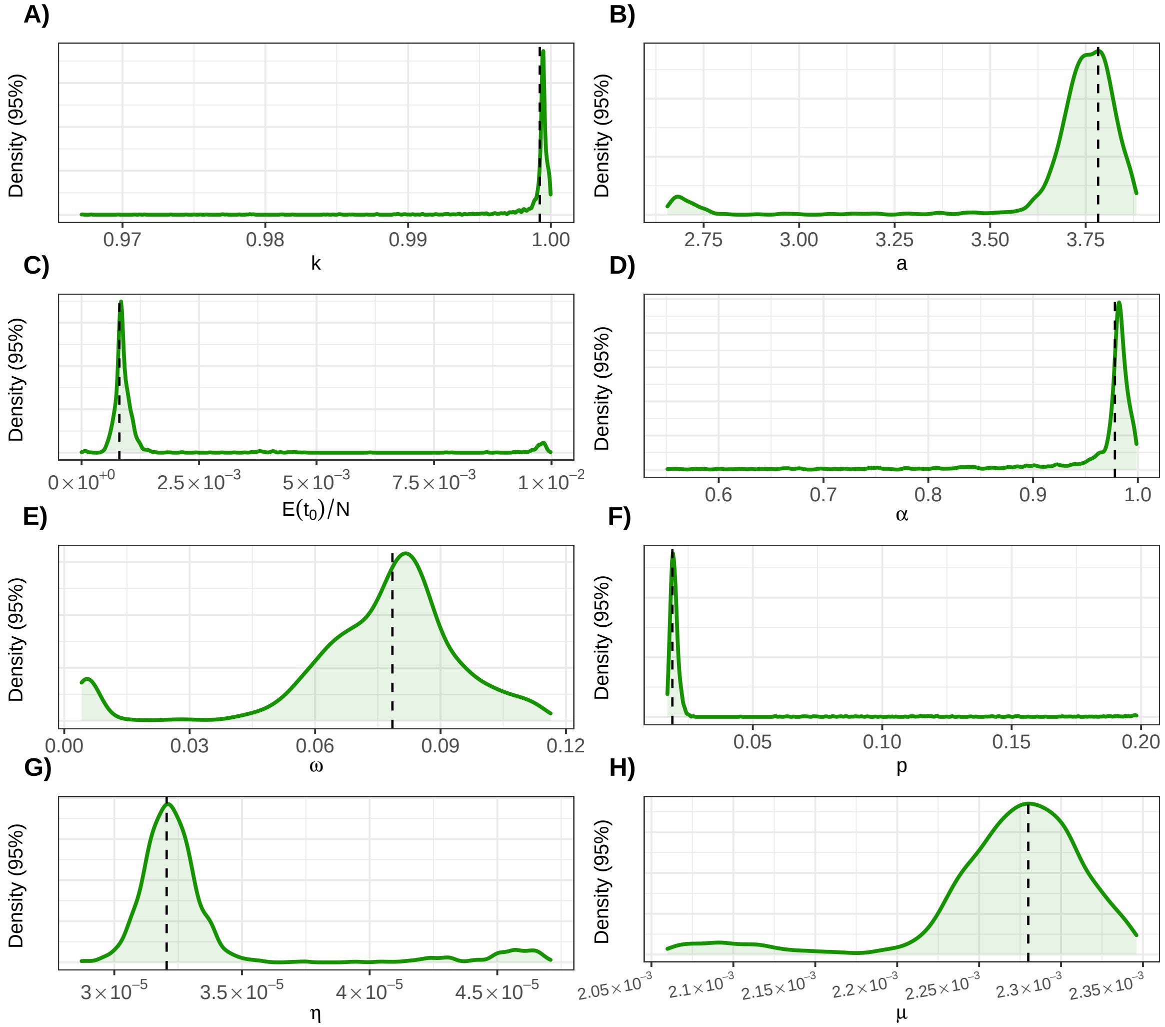


Figure S4: **95% distributions of the parameters estimated in the first step.** The distributions of these parameters (see Table S1 for definitions) were computed using wild bootstrap [8, 5]; *bootstraped* data were generated with residuals perturbed by an i.i.d. sequence of $n$ random weights $\{W_i\}_{i=1}^n$ following Mammen's 2-points distribution (that is, $(1-\sqrt{5})/2$ with probability $(\sqrt{5}+1)/(2\sqrt{5})$ and $(1+\sqrt{5})/2$ with probability $(\sqrt{5}-1)/(2\sqrt{5})$), which satisfies $\mathbb{E}(W_i) = 0$ and $\mathbb{E}(W_i^2) = 1$. We only represented values between the 2.5% quantile and the 97.5% quantile. The vertical dashed lines indicate the best (minimum) WLS estimates computed from the original data. Using Rademacher distribution (that is 1 or $-1$ equiprobably) instead yields very similar results (not shown).

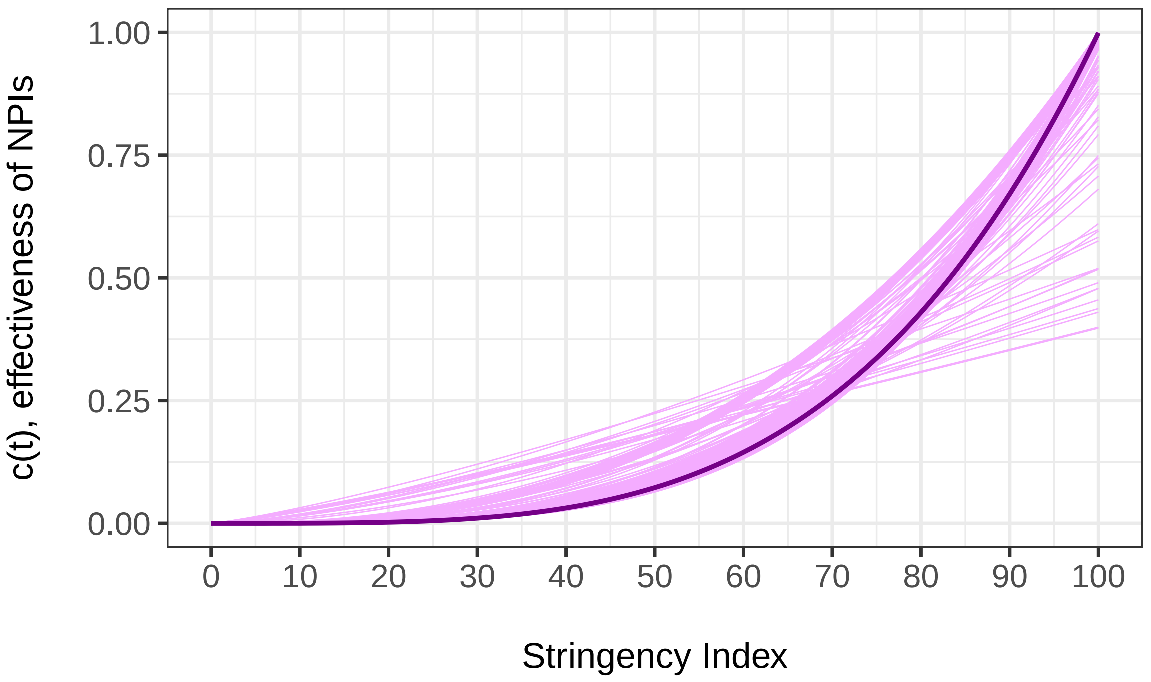


Figure S5: **Inferred relationship between the Stringency Index and the effectiveness of NPIs in the UK.** The link between the Stringency Index $\psi(t)$ and the effectiveness of NPIs $c(t)$ is modeled through the following function: $c(t) = k\,(\psi(t)/100)^a$. Depending on the value of $a$, this relationship may be concave ($0 < a < 1$), linear ($a = 1$) or convex ($a > 1$). The aim of the first step of our analysis is to infer the value of parameters $k$ and $a$. The best WLS estimator yielded $k = 1$ and $a = 3.78$ (dark line); a joint distribution for these two parameters (light lines) was obtained using wild bootstrap computations. With all our estimates of $a$ greater than 1, we always find a (more or less pronounced) convex relationship between the Stringency Index and the effectiveness of NPIs in the UK.

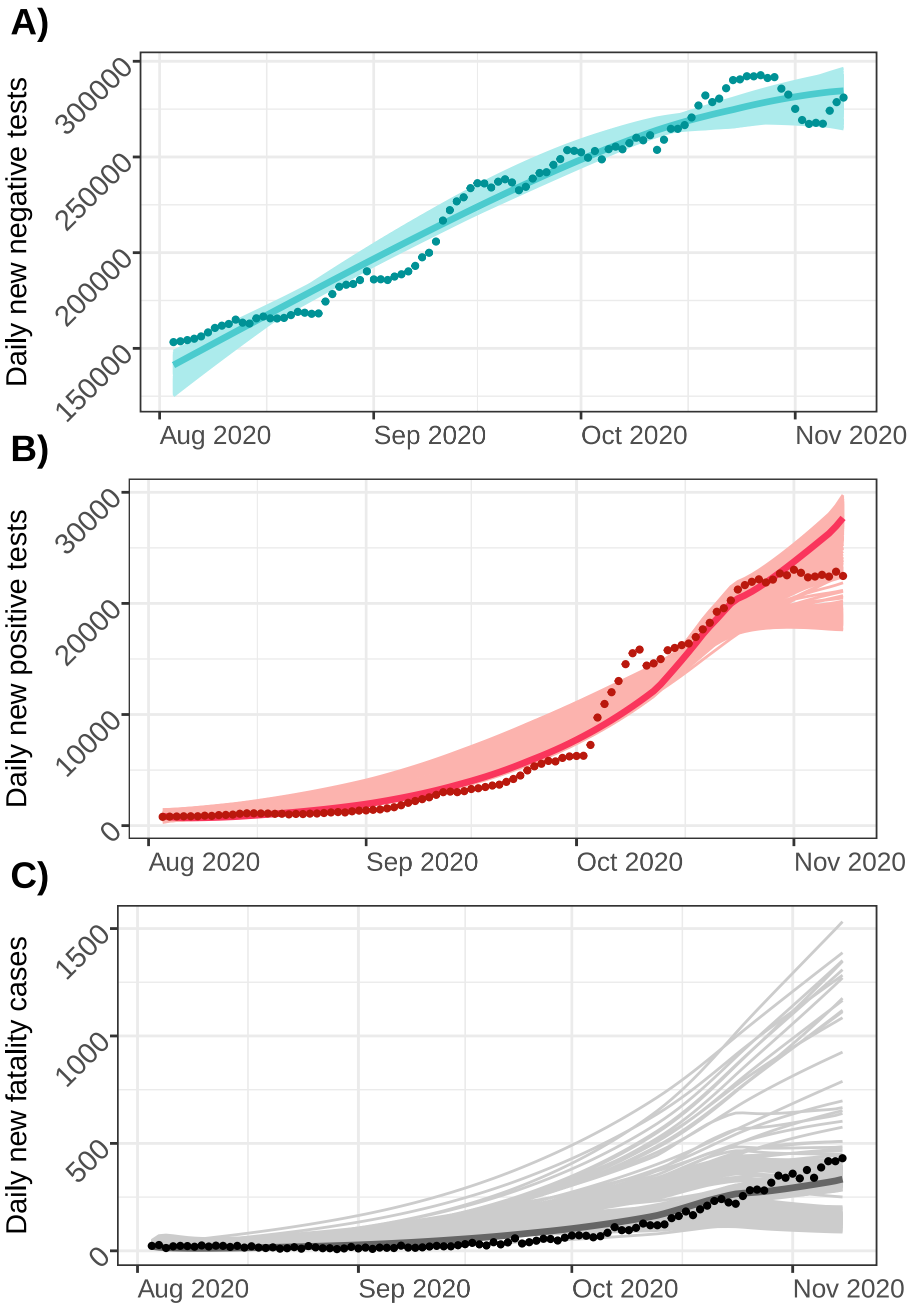


Figure S6: **Model fitting to data for the first phase (from 2020-08-03 to 2020-11-08 in the UK).** Observed data – (**A**) daily new cases tested negative, (**B**) daily new cases tested positive and (**C**) daily new fatality cases – are shown as dark points. Fits based on the best WLS estimates are represented as dark lines and about 2000 model fits resulting from wild bootstrap are represented as light lines.

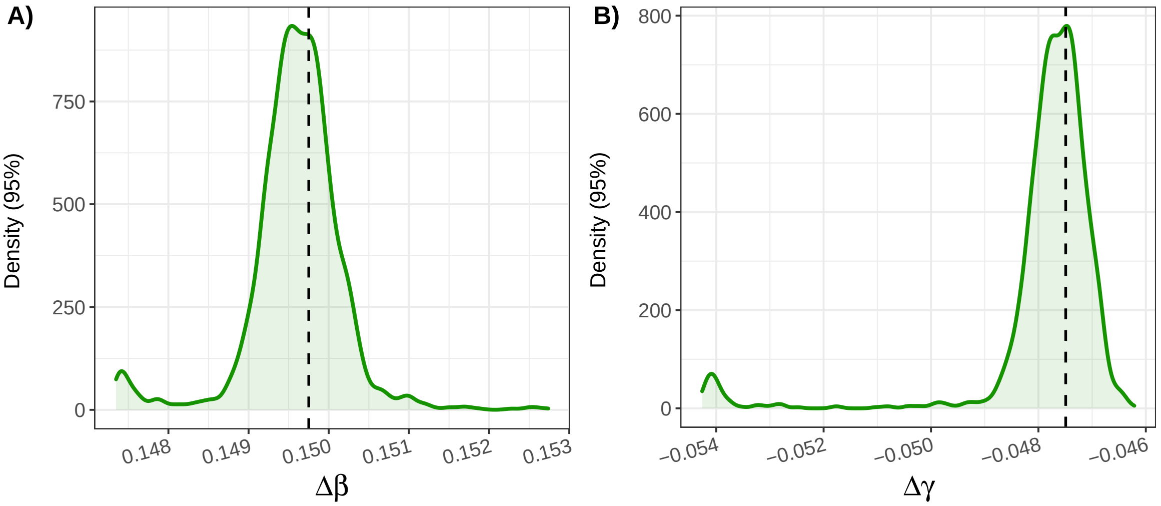


Figure S7: **95% distributions of the inferred phenotypic differences $\Delta\beta$ (transmission effect) and $\Delta\gamma$ (recovery effect) for the Alpha variant relative to the previous lineage.** Each value (per day) of $\Delta\beta$ and $\Delta\gamma$ was computed using a linear MEM and a particular pair $\{k; a\}$ that was previously estimated with wild bootstrap in the first step. Only values between the 2.5% quantile and the 97.5% quantile are represented. Vertical dashed lines indicate the estimates of $\Delta\beta$ and $\Delta\gamma$ using the best WLS estimates for parameters $k$ and $a$.

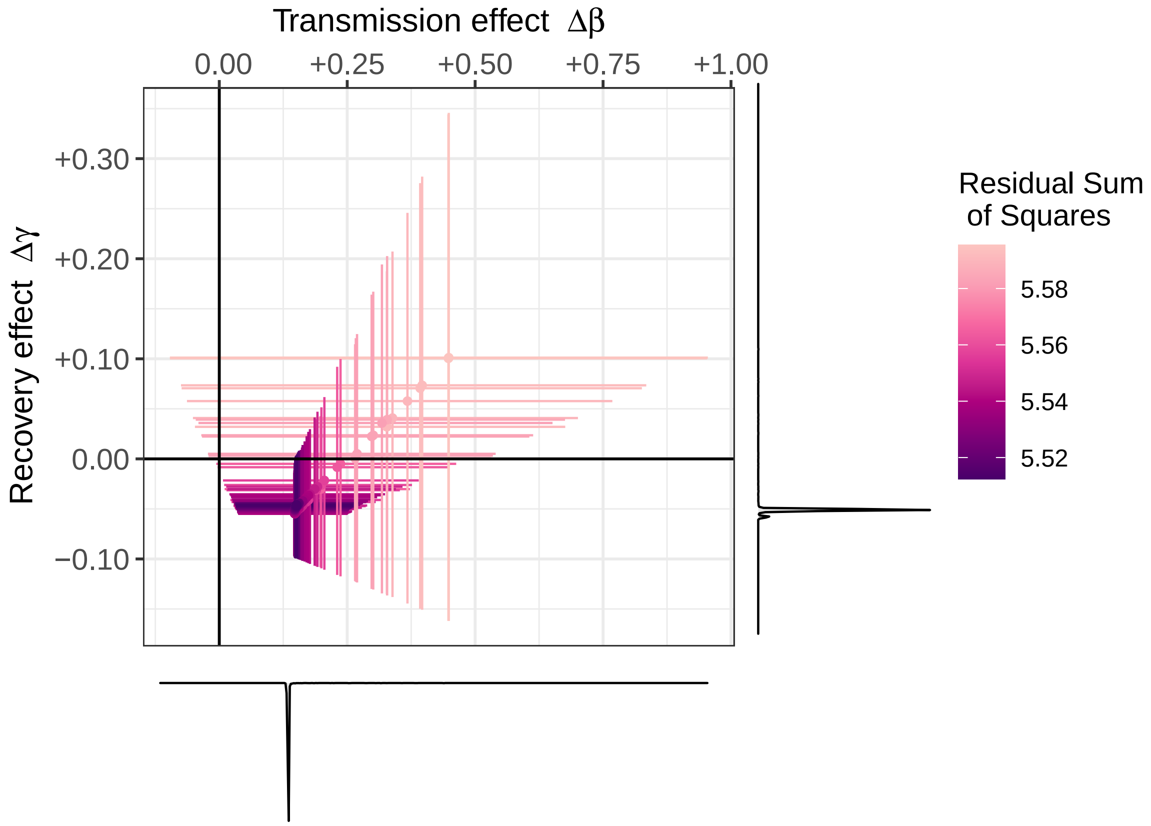


Figure S8: **Phenotypic profile of the Alpha variant (transmission and recovery rates) relative to the previous lineage with uncertainty on the control parameters propagated from the first step.** Estimates per day (points) and 95% confidence interval (crosses) are colored according to the residual sum of squares. We set $S/N = 0.75$, $\kappa = 0.2$, $\beta_w = 0.25$ and $\gamma_w = 0.1$ and each of the almost 2000 points corresponds to a pair $\{k; a\}$ that was previously estimated with wild bootstrap in the first step. Side curbs, representing the densities of the estimates of parameters $\Delta\beta$ (bottom) and $\Delta\gamma$ (right), show that the vast majority of these estimates are grouped around very similar values (dark purple). Indeed, for $\Delta\beta$, 95% of them are between 0.147 and 0.153, for which each corresponding 95% CI remain positive, while, for $\Delta\gamma$, 95% are between -0.054 and -0.046, among which 61% of the corresponding 95% CIs cross the zero axis. With these estimates of $\Delta\beta$ and $\Delta\gamma$, the selection coefficient $s(t)$ of the Alpha variant in the absence of NPI was computed, on average, around 0.11 per day (standard deviation: 0.003), that is 0.77 per week (standard deviation: 0.023).

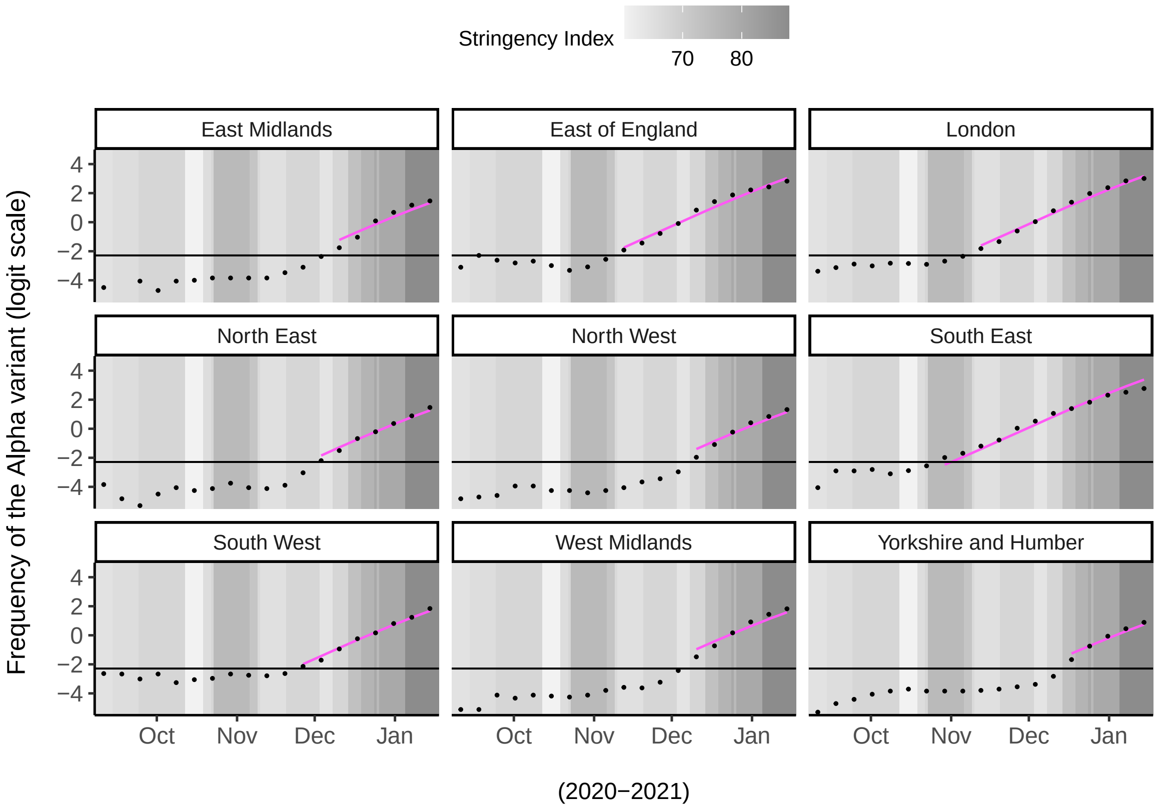


Figure S9: **Model fitting to logit-frequencies of the Alpha variant in the nine regions of England.** Frequencies of SGTF (black points, on logit scale) were used as a proxy for the Alpha variant. The magenta curbs show the fitted values based on a linear MEM with $\Delta\beta$ and $\Delta\gamma$ as fixed effects and the region as a random effect on the intercept of the model. We only fitted frequencies greater than or equal to the threshold frequency 10% (horizontal black lines) in order to get rid of the more stochastic part of these temporal dynamics (when the variant was not yet really well established in the host population).

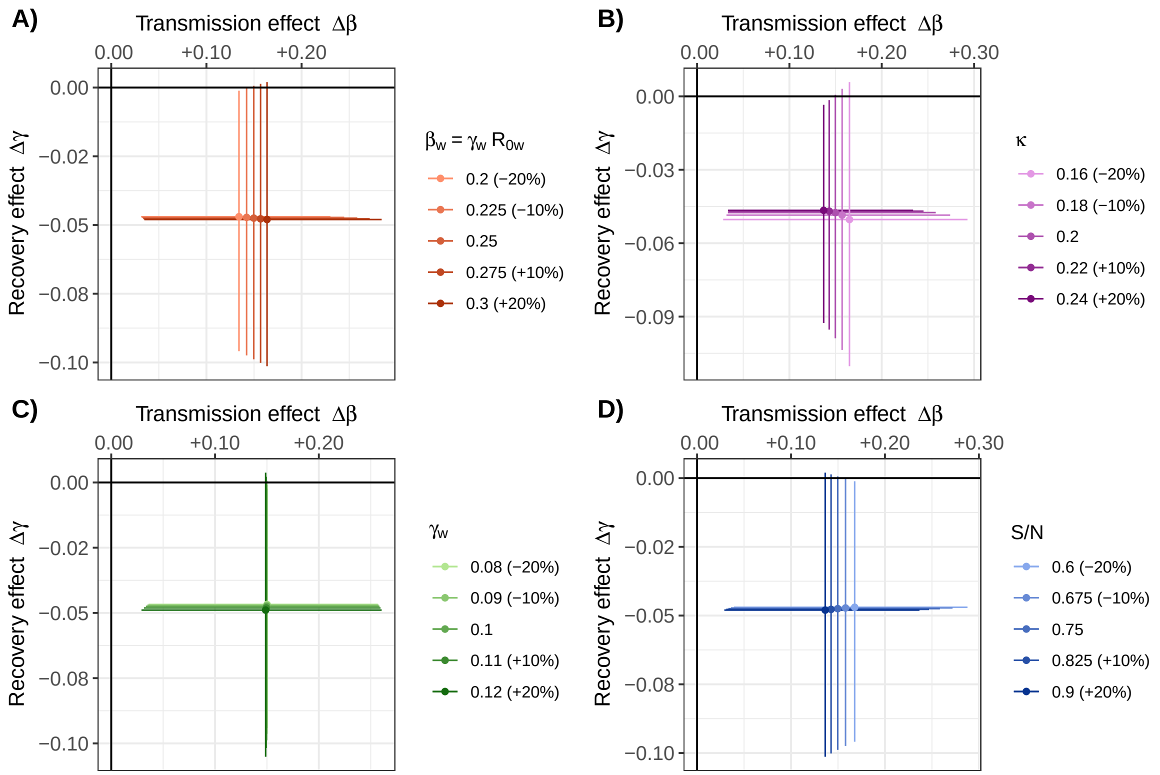


Figure S10: **Phenotypic differences between the Alpha variant and the resident strain with small variations in the fixed parameters.** $\pm 10\%$ and $\pm 20\%$ perturbations were applied separately to each fixed parameter to investigate robustness: $\beta_w = 0.25$ (A), $\kappa = 0.2$ (B), $\gamma_w = 0.1$ (C) and $S/N = 0.75$ (D). Keeping our best WLS estimates for control parameters $k$ and $a$ ($k = 1$ and $a = 3.78$), we reiterated MEMs to obtain new estimates (points) and 95% CIs (segments) of the phenotypic differences $\Delta\beta$ – transmission effect – and $\Delta\gamma$ – recovery effect – between the Alpha variant and the resident strain.

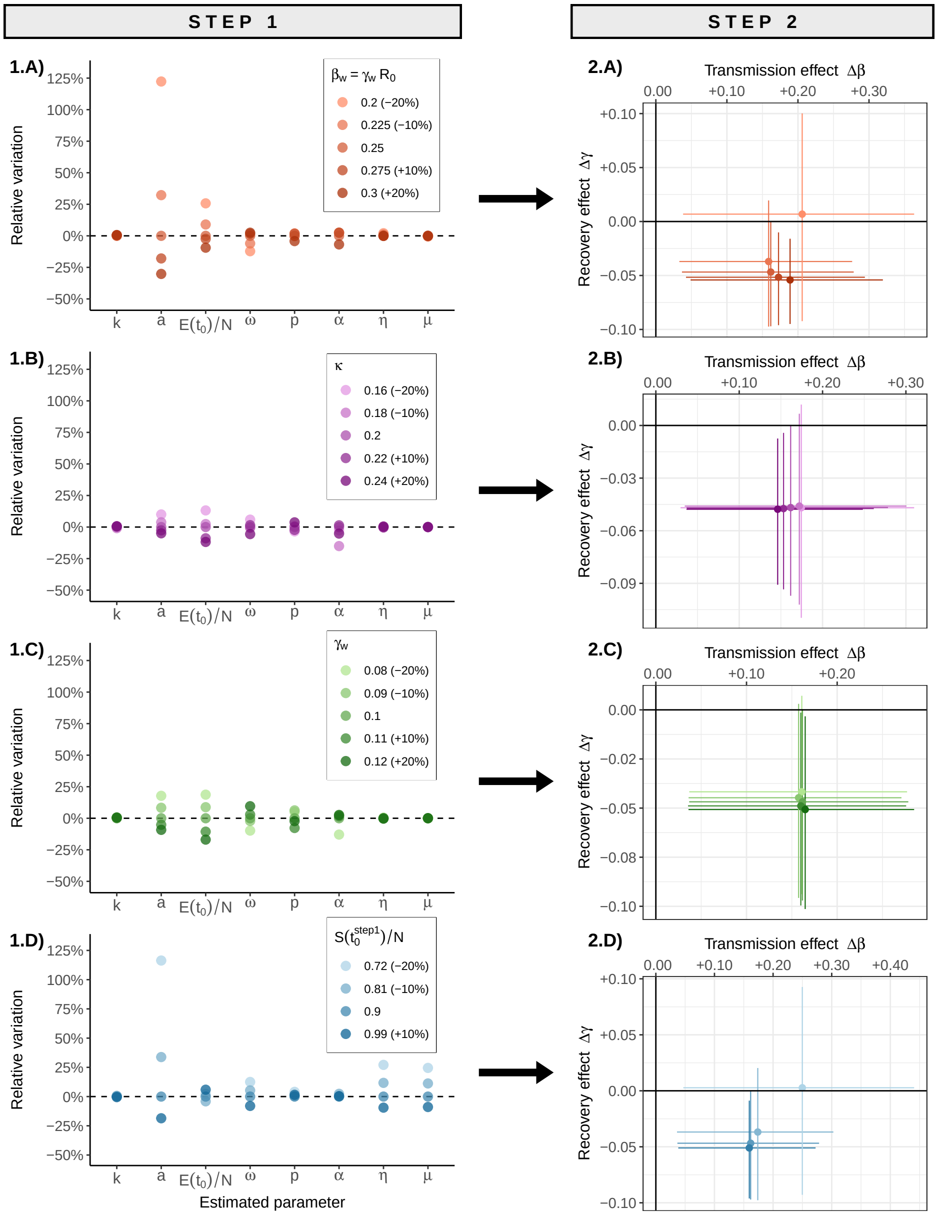


Figure S11: **Effects of small variations in the fixed parameters propagated through the two-step approach.** ±10% and ±20% perturbations were applied separately to each fixed parameter: (A) $\beta_w = 0.25$ (transmission rate of the WT), (B) $\kappa = 0.2$ (transition rate from $E$ (exposed state) to $I$ (infectious state); same for both strains), (C) $\gamma_w = 0.1$ (recovery rate of the WT) and (D) $S(t_0^{\text{step 1}})/N = 0.9$ (initial proportion of susceptible hosts where $t_0^{\text{step 1}}$ refers to the initial time point in step 1). (1) In the first step, each point corresponds to the best estimation (lowest WLS value) from 500 non-linear optimizations starting from uniformly drawn initial conditions (cf. **Table S2**); relative variations (y-axis) refer to the percentage of variation between the parameters estimated with perturbations and those without. (2) For the second step, we reiterated MEMs using estimates from (1) for the control parameters $k$ and $a$ along with the same corresponding ±10% and ±20% perturbations to obtain new estimates (points) and 95% CIs (segments) for the phenotypic differences $\Delta\beta$ and $\Delta\gamma$ ($S/N$ was set in (2) consistently with the end of each simulation in (1)).

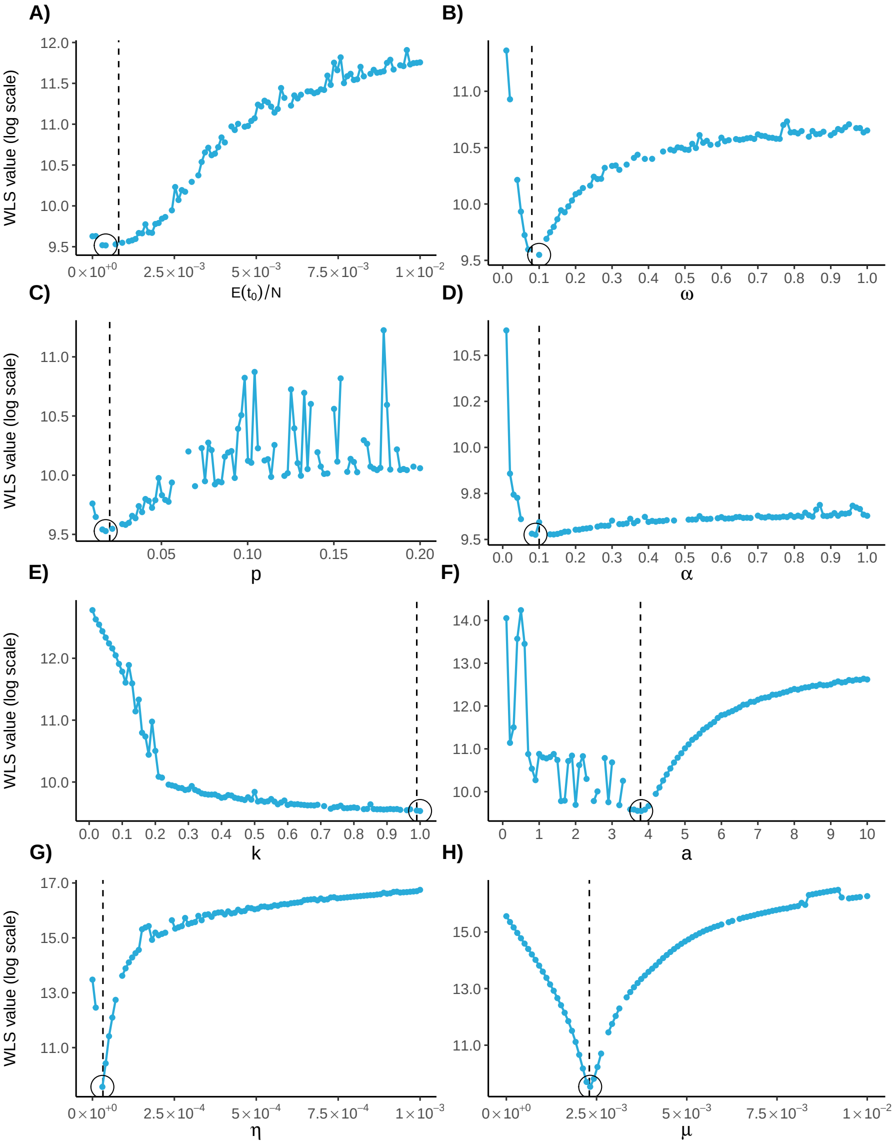


Figure S12: **Identifiability profiles of the first phase.** Simulated data for the first phase – i.e. (i) new cases tested negative, (ii) new cases tested positive and (iii) new fatality cases – were generated with the parameter values indicated by the vertical dashed lines: $E(t_0)/N = 8.0 \times 10^{-4}$ (where $t_0$ refers to the initial time point of the first phase), $\omega = 0.08$, $p = 0.02$, $\alpha = 0.1$, $k = 0.99$, $a = 3.78$, $\eta = 3.2 \times 10^{-5}$, $\mu = 2.3 \times 10^{-3}$; i.i.d. Gaussian noise was added to each series of simulated data, with standard deviation (i) 5000, (ii) 500 and (iii) 5. For the fixed parameters, we set: $\kappa = 0.2$, $\gamma = 0.1$, $\mathcal{R}_0 = 2.5$, $\beta = \gamma\mathcal{R}_0 = 0.25$ and $S(t_0)/N = 0.9$. We used real values for the Stringency Index (from 2020-08-02 to 2020-11-08 in the UK). Profiles were built following [9]: a parameter of interest is set to a given value (on the x-axis) and the others are estimated to obtain a WLS value (y-axis, here on log scale); this is then reiterated with different values of the parameter of interest to cover the desired range (x-axis). As initial conditions, we only started from the parameter values that we used to simulate the data. Discontinuities in the profiles result merely from convergence failures. Points associated with the lowest WLS value are enclosed in a circle and their good match with the values used to simulate the data (vertical dashed lines) confirms that these parameters should be identifiable. Though, the shallowness of some profiles suggest that precise estimations may be numerically difficult.

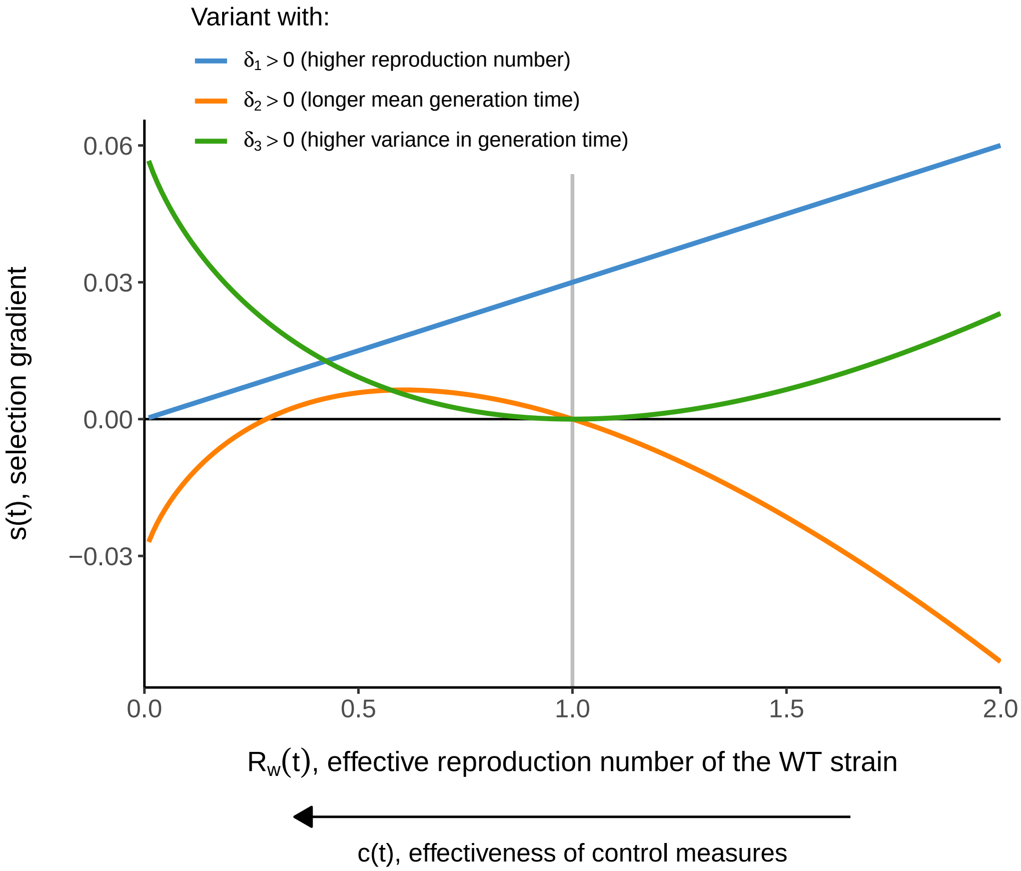


Figure S13: **Variation of the selection gradient of three types of variant as a function of the effective reproduction number of the resident strain (WT).** In [1], Blanquart *et al.* propose that an emerging variant $m$ may differ phenotypically from the WT strain $w$ by its effective reproduction number $\mathcal{R}_m(t) = \mathcal{R}_w(t)\big(1+\delta_1\big)$ and/or by its mean generation time $\mu_m = \mu_w(t)\big(1+\delta_2\big)$ and/or by the variance of its generation time $\sigma_m^2 = \big(\sigma_w(t)\big(1+\delta_3\big)\big)^2$ (we keep the same notations as in the original article [1] for the phenotypic differences $\delta_1$, $\delta_2$ and $\delta_3$). Generation times are assumed to be gamma-distributed and, in particular, exponentially distributed for the resident strain (special case of gamma distribution where $\sigma_w = \mu_w$) with $\mu_w = 10$. The selection gradient is computed under the assumption of weak selection and, for each curb, phenotypic differences are: $\big(\delta_1 = +30\%,\ \delta_2 = 0,\ \delta_3 = 0\big)$, $\big(\delta_1 = 0,\ \delta_2 = +30\%,\ \delta_3 = 0\big)$ and $\big(\delta_1 = 0,\ \delta_2 = 0,\ \delta_3 = +30\%\big)$, respectively. The variant is selected when its selection gradient is positive – conversely, counter-selected when it is negative. The case $\mathcal{R}_w(t) = 1$ (vertical grey line) corresponds to a stable epidemic. At the bottom of the figure, the horizontal arrow pointing to the left symbolizes more explicitly the impact of NPIs (control measures) that reduces $\mathcal{R}_w(t)$, therefore altering the selection gradient.

Table S1: **Summary of the parameters involved in the model of the first phase.** The first phase is the period that took place just before the emergence of the Alpha variant. $t_0^{\text{step 1}}$ refers to the time point at which the model is initialized.

| **Symbol** | **Description** | **Value** |
|---|---|---|
| $k$ | Maximum achievable efficacy of NPIs | estimated |
| $a$ | '*Shape*' parameter for the relationship between the efficacy of NPIs and the Stringency Index | estimated |
| $E(t_0^{\text{step 1}})$ | Initial number of exposed hosts | estimated |
| $\alpha$ | Virulence (*per capita* disease-induced mortality rate) | estimated |
| $\omega$ | Probability of symptom development | estimated |
| $p$ | Probability to die for symptomatic hosts | estimated |
| $\eta$ | Slope of the increase in screening effort over time | estimated |
| $\mu$ | Intercept of the increase in screening effort over time | estimated |
| $\mathcal{R}_0$ | Basic reproduction number | 2.5 [4, 6, 7] |
| $\gamma$ | *Per capita* recovery rate | 0.1 day$^{-1}$ [2] |
| $\beta$ | *Per capita* transmission rate | $\gamma\mathcal{R}_0 = 0.25$ day$^{-1}$ |
| $\kappa$ | Transition rate from exposed to infectious state | 0.2 day$^{-1}$ [3] |
| $N$ | 2020 UK population size | $\approx$ 67.9 million |
| $S(t_0^{\text{step 1}})/N$ | Initial proportion of susceptible indivuals | 0.9 |
| $I_{Sd}(t_0^{\text{step 1}})$ | Initial number of symptomatic hosts that will eventually die | $\frac{D(t_0^{\text{step 1}}+1)-D(t_0^{\text{step 1}})}{\alpha}$ |
| $I_{Sr}(t_0^{\text{step 1}})$ | Initial number of symptomatic hosts that will eventually recover | $\left(\frac{1-p}{p}\right) I_{Sd}(t_0^{\text{step 1}})$ |
| $I_A(t_0^{\text{step 1}})$ | Initial number of asymptomatic individual | $\left(\frac{1-\omega}{\omega}\right) I_S(t_0^{\text{step 1}})$ |
| $R(t_0^{\text{step 1}})$ | Initial number of recovered (immune) hosts | $N - S(t_0^{\text{step 1}}) - E(t_0^{\text{step 1}}) - I(t_0^{\text{step 1}})$ |

Table S2: **Summary of the initialization and optimization sets for the parameters estimated in the model of the first phase.** Because of the presence of local minima, optimization procedure was repeated for 1500 sets of initial values by drawing randomly in each initialization interval below according to a continuous uniform distribution. Parameter transformations enabled then to restrict optimization searches in more relevant ranges of values (referred to '*optimization intervals*' below). You may refer to the **Table S1** for the meaning of the symbols.

| Symbol | Initialization interval | Optimization interval |
|---|---|---|
| $k$ | $[0; 1]$ | $[0; 1]$ |
| $a$ | $[0; 10]$ | $\mathbb{R}_+^*$ |
| $E(t_0^{\text{step 1}})$ | $[0; 10^{-2}]$ | $[0; 10^{-2}]$ |
| $\alpha$ | $[0; 1]$ | $[0; 1]$ |
| $\omega$ | $[0.2; 0.8]$ | $[0; 1]$ |
| $p$ | $[0; 0.1]$ | $[0; 0.2]$ |
| $\eta$ | $[0; 10^{-4}]$ | $[0; 1]$ |
| $\mu$ | $[0; 10^{-2}]$ | $[0; 1]$ |

Table S3: **Summary of the parameters involved in the model of the second phase.** The second phase is the period that took place just after the emergence of the Alpha variant.

| Symbol | Description | Value |
|---|---|---|
| $\Delta\beta$ | Phenotypic difference between the Alpha variant and the resident strain in terms of transmission rate | estimated |
| $\Delta\gamma$ | Phenotypic difference between the Alpha variant and the resident strain in terms of recovery rate | estimated |
| $k$ | Maximum achievable efficacy of NPIs | estimates from the first step (best WLS estimate: 1) |
| $a$ | '*Shape*' parameter for the relationship between the efficacy of NPIs and the Stringency Index | estimates from the first step (best WLS estimate: 3.78) |
| $\gamma_w$ | *Per capita* recovery rate of the resident strain | 0.1 day$^{-1}$ [2] |
| $\beta_w$ | *Per capita* transmission rate of the resident strain | 0.25 day$^{-1}$ |
| $\kappa$ | Transition rate from exposed to infectious state | 0.2 day$^{-1}$ [3] |
| $S/N$ | Proportion of susceptible host (assumed constant for short enough periods of time during a controlled epidemic) | 0.75 |

Table S4: **Likelihood-based comparisons of nested linear MEM**. The tilde operator $\sim$ refers to the linear relationship between the response variable $\text{logit}(\widetilde{f}_m(t))$, the logit-frequency of the variant, and the explanatory variables. The phenotypic differences between the variant and the resident strain $\Delta\beta$ (transmission effect) and $\Delta\gamma$ (recovery effect) are considered as fixed effects while $(1|Region)$ refers to a random effect of the region on the intercept of the model. With a significance level of 5%, results show a significant effect (*) for $\Delta\beta$ but not (.) for $\Delta\gamma$ (although the *p-value* associated with the latter is very close to the significance threshold). AIC is the Akaike Information Criterion and BIC is the Bayesian Information Criterion (models with lower values are preferred).

| | | | $\text{logit}(\widetilde{f}_m(t)) \sim \Delta\beta + \Delta\gamma + (1\|Region)$ |
|---|---|---|---|
| | AIC | BIC | AIC 61.122 BIC 72.969 |
| $\text{logit}(\widetilde{f}_m(t)) \sim \Delta\beta + (1\|Region)$ | 62.867 | 72.345 | *p-value* $= 0.053$ (.) |
| $\text{logit}(\widetilde{f}_m(t)) \sim \Delta\gamma + (1\|Region)$ | 65.386 | 74.864 | *p-value* $= 0.012$ $(*)$ |
