## Supplementary material for "Phenotypic evolution of SARS-CoV-2: a statistical inference approach": SI Appendix

March 27, 2023

#### Preamble

In this appendix, we show how phenotypic traits of the new variant affect its temporal dynamics. We derive the selection coefficient (or selection gradient) of a variant – a measure of how much it is favoured or disfavoured through natural selection – in a susceptible-exposed-infectious-recovered ($SEIR$) model. In sections S2-S5 we show how we can use a weak selection argument to obtain a useful approximation of the selection coefficient of the new variant. In section S6 we use a weak selection argument to derive an approximation of the differentiation of variant frequency between the exposed and the infectious states. Finally, in section S7, we detail how our analysis relates to the approach used in [1].

#### S1 $SEIR$ model

Let us consider a directly and horizontally transmitted disease and a host population of size $N$ for which individuals are either susceptible ($S$), exposed ($E$, infected but not yet infectious), infectious ($I$) or recovered ($R$). For a given state, for instance $S$, we denote $S(t)$, where $t$ is the current time, its density and $\dot{S}(t)$, its differentiation with respect to time. Demographic parameters (newborns, migration balance, natural mortality, ...) are neglected.

Let also consider a polymorphic pathogen population: the WT strain (resident), which will be represented with the subscript $w$, and the mutant strain (or variant), which will be represented with the subscript $m$ (we then neglect any occurrence of new mutations). Therefore, $E(t)$ and $I(t)$ can be respectively decomposed into: $E(t) = E_w(t) + E_m(t)$ and $I(t) = I_w(t) + I_m(t)$. The variant may differ phenotypically from the WT in its effective transmission rate $\beta_m = \beta_w + \Delta\beta$ and/or in its recovery rate $\gamma_m = \gamma_w + \Delta\gamma$ and/or in its disease-induced mortality rate (virulence) $\alpha_m = \alpha_w + \Delta\alpha$ and/or in its transition rate from state $E$ to state $I$ $\kappa_m = \kappa_w + \Delta\kappa$ (note that $1/\kappa$ is thus the mean sojourn time in the exposed state, i.e. the latent period).

Besides, the transmission of both strains is more or less affected depending on $c(t)$, the effectiveness of governmental control measures – i.e. Non-Pharmaceutical Interventions (NPIs) – implemented at time $t$ to mitigate the spread of the epidemic.

We also assume that superinfections do not occur – including co-infections with both strains – and that (persistent) immunity acquired with either strain protects effectively against both. We model the temporal dynamics of this $SEIR$ system with the following set of ordinary differential equations (ODEs):

$$\begin{cases} \dot{S}(t) = -(1-c(t))\overline{\beta}(t)S(t)\dfrac{I(t)}{N} \\ \dot{E}(t) = (1-c(t))\overline{\beta}(t)S(t)\dfrac{I(t)}{N} - \overline{\kappa}(t)E(t) \\ \dot{I}(t) = \overline{\kappa}(t)E(t) - \Big(\overline{\alpha}(t) + \overline{\gamma}(t)\Big)I(t) \\ \dot{R}(t) = \overline{\gamma}(t)I(t) \end{cases} \tag{S1}$$

The overlines refer to mean values of the phenotypic traits after averaging over the distribution of strain frequencies in the relevant compartments of the model:

$$\begin{cases} \overline{\kappa}(t) = (1-p_m(t))\kappa_w + p_m(t)\kappa_m \\ \overline{\beta}(t) = (1-q_m(t))\beta_w + q_m(t)\beta_m \\ \overline{\gamma}(t) = (1-q_m(t))\gamma_w + q_m(t)\gamma_m \\ \overline{\alpha}(t) = (1-q_m(t))\alpha_w + q_m(t)\alpha_m \end{cases}$$

where the frequency of the variant $m$ in the compartment $E$ is noted $p_m(t) = E_m(t)/E(t)$ and the frequency of the variant $m$ in the compartment $I$ is noted $q_m(t) = I_m(t)/I(t)$. For each strain $i$ ($i \in \{w; m\}$):

$$\begin{cases} \dot{E}_i(t) = (1-c(t))\beta_i S(t)\dfrac{I_i(t)}{N} - \kappa_i E_i(t) \\ \dot{I}_i(t) = \kappa_i E_i(t) - \Big(\alpha_i + \gamma_i\Big)I_i(t) \end{cases}$$

By noting $\mathbf{X}(t) = \begin{pmatrix} E(t) & I(t) \end{pmatrix}^\top$, we have:

$$\dot{\mathbf{X}}(t) = \overline{\mathbf{R}}(t)\mathbf{X}(t)$$

with $\overline{\mathbf{R}}(t) = \begin{pmatrix} -\overline{\kappa}(t) & (1-c(t))\overline{\beta}(t)\frac{S(t)}{N} \\ \overline{\kappa}(t) & -\overline{\alpha}(t) - \overline{\gamma}(t) \end{pmatrix}$, the matrix of (average) transitions rates.

### S2 Overall frequency of the variant

The overall frequency of the variant in the system at time $t$, $f_m(t)$, is:

$$f_m(t) = \frac{E_m(t) + I_m(t)}{E(t) + I(t)} = \frac{p_m(t)E(t) + q_m(t)I(t)}{E(t) + I(t)} = p_m(t)p(t) + q_m(t)q(t)$$

where $p(t) = E(t)/(E(t) + I(t))$ and $q(t) = I(t)/(E(t) + I(t))$ are the class frequencies of infected individuals in the exposed state $E$ and in the infectious state $I$, respectively. Note therefore that $p(t) + q(t) = 1$.

The temporal dynamics of the variant can be tracked more conveniently using the following quantity [4, 5]:

$$\widetilde{f}_m(t) = p_m(t)v^E(t)p(t) + q_m(t)v^I(t)q(t)$$

where, $v^E(t)$ and $v^I(t)$ are the reproductive values in state $E$ and in state $I$, respectively.
Let $\mathbf{v}(t) = \begin{pmatrix} v^E(t) & v^I(t) \end{pmatrix}^\top$ be the vector of reproductive values and $\mathbf{f}(t) = \begin{pmatrix} p(t) & q(t) \end{pmatrix}^\top$ be the vector of frequencies within infected states; these two vectors are co-normalized such that $\mathbf{v}^\top \mathbf{f} = 1$. Under the assumption of weak selection, $\widetilde{f}_m(t)$ yields indeed a really useful expression:

$$\frac{\mathrm{d}\widetilde{f}_m(t)}{\mathrm{d}t} = \underbrace{\widetilde{f}_m(t)(1 - \widetilde{f}_m(t))}_{\text{Genetic variance}} \; \underbrace{\mathbf{v}(t)^\top \Delta\mathbf{R}(t)\mathbf{f}(t)}_{\text{s(t), selection coefficient}} \; , \tag{S2}$$

or more simply, using the logit function, that is ln(frequency of the variant / frequency of the WT strain):

$$s(t) = \frac{\mathrm{d}\ \mathrm{logit}(\widetilde{f}_m(t))}{\mathrm{d}t} = \mathbf{v}(t)^\top \Delta\mathbf{R}(t)\mathbf{f}(t). \tag{S3}$$

$\Delta\mathbf{R}(t)$ is the matrix of differences in transition rates between the mutant and the WT strains such that:

$$\Delta\mathbf{R}(t) = \mathbf{R}_m(t) - \mathbf{R}_w(t) = \begin{pmatrix} -\Delta\kappa & (1 - c(t))\Delta\beta\frac{S(t)}{N} \\ \Delta\kappa & -\Delta\alpha - \Delta\gamma \end{pmatrix}.$$

The selection coefficient $s(t)$ is thus given by:

$$\begin{aligned} s(t) &= \mathbf{v}(t)^\top \Delta\mathbf{R}(t)\mathbf{f}(t) \\ &= p(t)\Delta\kappa\Big(v^I(t) - v^E(t)\Big) + q(t)\left[(1 - c(t))\Delta\beta\frac{S(t)}{N}v^E(t) - \Big(\Delta\alpha + \Delta\gamma\Big)v^I(t)\right]. \end{aligned} \tag{S4}$$

The main problem with this theoretical expression is that class frequencies ($p(t)$ and $q(t)$) and reproductive values ($v^E(t)$ and $v^I(t)$) are generally not available from public health data. In the following section we show how we can derive useful approximations for these quantities.

### S3 Class frequencies within infected states & growth rate

Following [4, 5], the temporal dynamics of the vector of class frequencies is given by:

$$\frac{\mathrm{d}\mathbf{f}(t)}{\mathrm{d}t} = \overline{\mathbf{R}}(t)\mathbf{f}(t) - \overline{r}(t)\mathbf{f}(t)$$

with $\overline{r}(t)$, the growth rate of the epidemic:

$$\overline{r}(t) \;=\; \mathbf{1}^\top \overline{\mathbf{R}}(t)\mathbf{f}(t) \;=\; q(t)\Big((1-c(t))\overline{\beta}(t)\frac{S(t)}{N} - \overline{\alpha}(t) - \overline{\gamma}(t)\Big). \tag{S5}$$

Therefore:

$$\begin{cases} \dot{p}(t) &= q(t)(1-c(t))\overline{\beta}(t)\dfrac{S(t)}{N} - p(t)\Big(\overline{\kappa}(t) + \overline{r}(t)\Big) \\ \dot{q}(t) &= p(t)\overline{\kappa}(t) - q(t)\Big(\overline{\alpha}(t) + \overline{\gamma}(t) + \overline{r}(t)\Big) \end{cases} \tag{S6}$$

When the difference between the vital rates of the mutant vs. resident is small and $\mathcal{O}(\varepsilon)$ (i.e. weak selection), the dynamics of the frequency $\tilde{f}_m(t)$ is also $\mathcal{O}(\varepsilon)$ (equation (S2)) while equation (S6) is $\mathcal{O}(1)$. This implies that $p(t)$ and $q(t)$ can be treated as fast variables while $\widetilde{f}_m(t)$ is a slow variable. Using a quasi-equilibrium approximation, i.e. setting the right-hand sides of (S6) to 0, we then have:

$$\begin{cases} q(t)(1-c(t))\overline{\beta}(t)\dfrac{S(t)}{N} &= p(t)\Big(\overline{\kappa}(t) + \overline{r}(t)\Big) \\ p(t)\overline{\kappa}(t) &= q(t)\Big(\overline{\alpha}(t) + \overline{\gamma}(t) + \overline{r}(t)\Big) \end{cases}$$

which yields:

$$\frac{p(t)}{q(t)} = \frac{(1-c(t))\overline{\beta}(t)\frac{S(t)}{N}}{\overline{\kappa}(t) + \overline{r}(t)} = \frac{\overline{\alpha}(t) + \overline{\gamma}(t) + \overline{r}(t)}{\overline{\kappa}(t)}. \tag{S7}$$

In addition,

$$\frac{p(t)}{q(t)} = \frac{\overline{\alpha}(t) + \overline{\gamma}(t) + \overline{r}(t)}{\overline{\kappa}(t)} \qquad \Longleftrightarrow \qquad \frac{1-q(t)}{q(t)} = \frac{\overline{\alpha}(t) + \overline{\gamma}(t) + q(t)\Big((1-c(t))\overline{\beta}(t)\frac{S(t)}{N} - \overline{\alpha}(t) - \overline{\gamma}(t)\Big)}{\overline{\kappa}(t)}$$

$$\Longleftrightarrow \qquad q(t)^2\Big((1-c(t))\overline{\beta}(t)\frac{S(t)}{N} - \overline{\alpha}(t) - \overline{\gamma}(t)\Big) + q(t)\Big(\overline{\kappa}(t) + \overline{\alpha}(t) + \overline{\gamma}(t)\Big) - \overline{\kappa}(t) = 0$$

Only the following solution satisfies $q(t) \in [0,1]$:

$$q(t) = \frac{-\Big(\overline{\kappa}(t) + \overline{\alpha}(t) + \overline{\gamma}(t)\Big) + \sqrt{\Big(\overline{\kappa}(t) - \overline{\alpha}(t) - \overline{\gamma}(t)\Big)^2 + 4\overline{\kappa}(t)(1-c(t))\overline{\beta}(t)\frac{S(t)}{N}}}{2\Big((1-c(t))\overline{\beta}(t)\frac{S(t)}{N} - \overline{\alpha}(t) - \overline{\gamma}(t)\Big)}$$

Under weak selection and when $q(t)$ is at equilibrium, the growth rate of the epidemic can thus be approximated by:

$$\overline{r}(t) \approx \frac{1}{2}\left[-\overline{\kappa}(t) - \overline{\alpha}(t) - \overline{\gamma}(t) + \sqrt{\Big(\overline{\kappa}(t) - \overline{\alpha}(t) - \overline{\gamma}(t)\Big)^2 + 4\overline{\kappa}(t)(1-c(t))\overline{\beta}(t)\frac{S(t)}{N}}\;\right] \tag{S8}$$

Note that, when $(1-c(t))\overline{\beta}(t)S(t)/N - \overline{\alpha}(t) - \overline{\gamma}(t) = 0$, then $\overline{r}(t) = 0$ which is also consistent for its approximation (S8).

Besides, starting from either the real expression of the growth rate (S5) or its approximation (S8), we have:

$$\lim_{\kappa \to +\infty} \overline{r}(t) = (1-c(t))\overline{\beta}(t)\frac{S(t)}{N} - \overline{\alpha}(t) - \overline{\gamma}(t).$$

In other words, we recover the growth rate of the corresponding nested $SIR$ model as a limit of this model.

### S4 Reproductive values within infected states

Following [4, 5], the temporal dynamics of reproductive values are given by:

$$\begin{aligned}
\frac{\mathrm{d}\mathbf{v}(t)^\top}{\mathrm{d}t} &= -\mathbf{v}(t)^\top \overline{\mathbf{R}}(t) + \overline{r}(t)\mathbf{v}(t)^\top \\
&= -\begin{pmatrix} v^E(t) & v^I(t) \end{pmatrix} \begin{pmatrix} -\overline{\kappa}(t) & (1-c(t))\overline{\beta}(t)\frac{S(t)}{N} \\ \overline{\kappa}(t) & -\overline{\alpha}(t) - \overline{\gamma}(t) \end{pmatrix} + \overline{r}(t)\begin{pmatrix} v^E(t) & v^I(t) \end{pmatrix}.
\end{aligned}$$

Therefore:

$$\begin{cases}
\dfrac{\mathrm{d}v^E(t)}{\mathrm{d}t} &= \overline{\kappa}(t)\Big(v^E(t) - v^I(t)\Big) + \overline{r}(t)v^E(t) \\
\dfrac{\mathrm{d}v^I(t)}{\mathrm{d}t} &= -(1-c(t))\overline{\beta}(t)\dfrac{S(t)}{N}v^E(t) + \Big(\overline{\alpha}(t) + \overline{\gamma}(t) + \overline{r}(t)\Big)v^I(t)
\end{cases} \tag{S9}$$

As previously, we see that the reproductive values are fast variables, so that we can use a quasi-equilibrium approximation. Setting the right-hand sides of (S9) become equal to 0, we obtain:

$$\begin{cases}
\overline{\kappa}(t)v^I(t) &= \Big(\overline{r}(t) + \overline{\kappa}(t)\Big)v^E(t) \\
(1-c(t))\overline{\beta}(t)\dfrac{S(t)}{N}v^E(t) &= \Big(\overline{\alpha}(t) + \overline{\gamma}(t) + \overline{r}(t)\Big)v^I(t)
\end{cases}$$

Which yields:

$$\frac{v^E(t)}{v^I(t)} = \frac{\overline{\kappa}(t)}{\overline{r}(t) + \overline{\kappa}(t)} = \frac{\overline{\alpha}(t) + \overline{\gamma}(t) + \overline{r}(t)}{(1-c(t))\overline{\beta}(t)\frac{S(t)}{N}} \tag{S10}$$

### S5 Approximation of the selection coefficient of the variant

Using the quasi-equilibrium approximation for vectors $\mathbf{f}(t)$ and $\mathbf{v}(t)$ (*cf.* (S7) and (S10), respectively), the expression of the selection coefficient in equation (S4) becomes:

$$s(t) = p(t)\Delta\kappa\left(v^E(t)\frac{\overline{r}(t)+\overline{\kappa}(t)}{\overline{\kappa}(t)} - v^E(t)\right) + q(t)\left[(1-c(t))\Delta\beta\frac{S(t)}{N}v^I(t)\frac{\overline{r}(t)+\overline{\alpha}(t)+\overline{\gamma}(t)}{(1-c(t))\overline{\beta}(t)\frac{S(t)}{N}} - \left(\Delta\alpha+\Delta\gamma\right)v^I(t)\right]$$

$$= p(t)v^E(t)\overline{r}(t)\frac{\Delta\kappa}{\overline{\kappa}(t)} + q(t)v^I(t)\left[\frac{\Delta\beta}{\overline{\beta}(t)}\left(\overline{r}(t)+\overline{\alpha}(t)+\overline{\gamma}(t)\right) - \Delta\alpha - \Delta\gamma\right].$$

Since combining quasi-equilibrium approximations (S7) and (S10) yields

$$\frac{p(t)v^E(t)}{q(t)v^I(t)} = \frac{\overline{\alpha}(t)+\overline{\gamma}(t)+\overline{r}(t)}{\overline{\kappa}(t)+\overline{r}(t)}$$

and, using the co-normalization $\mathbf{v}^\top\mathbf{f} = p(t)v^E(t)+q(t)v^I(t) = 1$, then:

$$q(t)v^I(t) = \frac{\overline{\kappa}(t)+\overline{r}(t)}{\overline{\kappa}(t)+\overline{\alpha}(t)+\overline{\gamma}(t)+2\overline{r}(t)} \qquad \text{and} \qquad p(t)v^E(t) = \frac{\overline{\alpha}(t)+\overline{\gamma}(t)+\overline{r}(t)}{\overline{\kappa}(t)+\overline{\alpha}(t)+\overline{\gamma}(t)+2\overline{r}(t)}.$$

Thus:

$$s(t) = \frac{\left(\overline{\alpha}(t)+\overline{\gamma}(t)+\overline{r}(t)\right)\overline{r}(t)\frac{\Delta\kappa}{\overline{\kappa}(t)} + \left(\overline{\kappa}(t)+\overline{r}(t)\right)\left[\frac{\Delta\beta}{\overline{\beta}(t)}\left(\overline{r}(t)+\overline{\alpha}(t)+\overline{\gamma}(t)\right) - \Delta\alpha-\Delta\gamma\right]}{\overline{\kappa}(t)+\overline{\alpha}(t)+\overline{\gamma}(t)+2\overline{r}(t)} \qquad \text{(S11)}$$

Using (S8) to approximate the growth rate of the epidemic, the selection coefficient of the variant becomes after some rearrangements:

$$s(t) \approx \frac{2(1-c(t))\frac{S(t)}{N}\left(\Delta\kappa\overline{\beta}(t)+\overline{\kappa}(t)\Delta\beta\right) + \Delta\kappa\left(\overline{\kappa}(t)-\overline{\alpha}(t)-\overline{\gamma}(t)-Z(t)\right) - \left(\Delta\alpha+\Delta\gamma\right)\left(\overline{\kappa}(t)-\overline{\alpha}(t)-\overline{\gamma}(t)+Z(t)\right)}{2Z(t)} \qquad \text{(S12)}$$

with

$$Z(t) = \sqrt{\left(\overline{\kappa}(t)-\overline{\alpha}(t)-\overline{\gamma}(t)\right)^2 + 4\overline{\kappa}(t)(1-c(t))\overline{\beta}(t)\frac{S(t)}{N}}.$$

Here again, we can recover the expression of $s(t)$ from the nested $SIR$ model [3, 2] by taking the limit:

$$\lim_{\kappa \to +\infty} s(t) \quad = \quad (1-c(t))\Delta\beta\frac{S(t)}{N} - \Delta\alpha - \Delta\gamma.$$

As in the main text, we now assume that the virulence may be neglected ($\alpha_m = \alpha_w = 0$) and that there is no difference between the variant and the WT strains in terms of latent period, i.e. $\Delta\kappa = 0$.

The approximation (S11) of the selection coefficient reduces then to:

$$s(t) = \frac{\kappa + \overline{r}(t)}{\kappa + \overline{\gamma}(t) + 2\overline{r}(t)} \left[ \frac{\Delta\beta}{\overline{\beta}(t)} \left( \overline{r}(t) + \overline{\gamma}(t) \right) - \Delta\gamma \right] \tag{S13}$$

Or, using (S12), i.e. based on an approximation of the growth rate:

$$s(t) \approx \frac{2\kappa(1-c(t))\Delta\beta\frac{S(t)}{N} - \left(\kappa - \overline{\gamma}(t)\right)\Delta\gamma - \sqrt{\left(\kappa - \overline{\gamma}(t)\right)^2 + 4\kappa(1-c(t))\overline{\beta}(t)\frac{S(t)}{N}}\ \Delta\gamma}{2\ \sqrt{\left(\kappa - \overline{\gamma}(t)\right)^2 + 4\kappa(1-c(t))\overline{\beta}(t)\frac{S(t)}{N}}} \tag{S14}$$

Since d $\text{logit}(\widetilde{f}_m(t))/\text{d}t = s(t)$, we get the following approximation of $\text{logit}(\widetilde{f}_m(t))$ by integrating the last approximation of $s(t)$ between the time points $t_0$ and $t = t_0 + \Delta t$:

$$\begin{aligned} \text{logit}(\widetilde{f}_m(t)) \approx \text{logit}(\widetilde{f}_m(t_0)) + \kappa \int_{t_0}^{t} \left( \frac{(1-c(t))S(t)/N}{\sqrt{(\kappa - \overline{\gamma}(t))^2 + 4\kappa(1-c(t))\overline{\beta}(t)S(t)/N}} \right) dt\ \ \Delta\beta \\ - \frac{1}{2} \left[ \int_{t_0}^{t} \left( \frac{\kappa - \overline{\gamma}(t)}{\sqrt{(\kappa - \overline{\gamma}(t))^2 + 4\kappa(1-c(t))\overline{\beta}(t)S(t)/N}} \right) dt + \Delta t \right] \Delta\gamma \end{aligned} \tag{S15}$$

Assuming also that $\overline{\gamma}(t) \approx \gamma_w$, $\overline{\beta}(t) \approx \beta_w$ (weak selection) and $S(t)/N \approx S/N$ – i.e. the proportion of susceptible individuals varies sufficiently little throughout the considered time period –, we eventually obtain the expression we used in the main text:

$$\begin{aligned} \text{logit}(\widetilde{f}_m(t)) \approx \text{logit}(\widetilde{f}_m(t_0)) + \kappa \int_{t_0}^{t} \left( \frac{(1-c(t))}{\sqrt{(\kappa - \gamma_w)^2 + 4\kappa(1-c(t))\beta_w S/N}} \right) dt\ \ \Delta\beta \frac{S}{N} \\ - \frac{1}{2} \left[ \left(\kappa - \gamma_w\right) \int_{t_0}^{t} \left( \frac{1}{\sqrt{(\kappa - \gamma_w)^2 + 4\kappa(1-c(t))\beta_w S/N}} \right) dt + \Delta t \right] \Delta\gamma \end{aligned} \tag{S16}$$

### S6 Differentiation between the exposed and the infectious compartments

In this section, we start with a $SEIR$ model in a very general form. The particular transition rates used previously and in the main text will be specified after the study of this general case.

By taking up the matrix form of the temporal dynamics of $\mathbf{X}(t) = \begin{pmatrix} E(t) & I(t) \end{pmatrix}^\top$:

$$\dot{\mathbf{X}}(t) = \overline{\mathbf{R}}(t)\mathbf{X}(t)$$

with $\overline{\mathbf{R}}(t) = \begin{pmatrix} \overline{r}^{\overleftarrow{EE}} & \overline{r}^{\overleftarrow{EI}} \\ \overline{r}^{\overleftarrow{IE}} & \overline{r}^{\overleftarrow{II}} \end{pmatrix}$, the matrix of average transitions rates (the arrows indicate the sense of the transitions). We recall that the overlines refer to mean values of the phenotypic traits after averaging over the distribution of strain frequencies, such that:

$$\begin{cases} \overline{r}^{\overleftarrow{EE}} = r_m^{\overleftarrow{EE}} - (1-p_m(t))\Delta r^{\overleftarrow{EE}} \\ \overline{r}^{\overleftarrow{EI}} = r_m^{\overleftarrow{EI}} - (1-q_m(t))\Delta r^{\overleftarrow{EI}} \\ \overline{r}^{\overleftarrow{IE}} = r_m^{\overleftarrow{IE}} - (1-p_m(t))\Delta r^{\overleftarrow{IE}} \\ \overline{r}^{\overleftarrow{II}} = r_m^{\overleftarrow{II}} - (1-q_m(t))\Delta r^{\overleftarrow{II}} \end{cases}$$

In which, with $(i,j) \in \{E,I\}^2$, we denote the phenotypic differences: $\Delta r^{\overleftarrow{ji}} = r_m^{\overleftarrow{ji}} - r_w^{\overleftarrow{ji}}$, where the subscript $m$ refer to the variant and the subscript $w$ to the WT.

Note that these transition rates may or may not be time-dependent, depending on the model. For the sake of simplicity, we do not make here this (potential) time dependency explicit.

The temporal dynamics of $p_m(t)$ and $q_m(t)$ are given by:

$$\begin{cases} \dot{p}_m(t) = & p_m(t)(1-p_m(t))\Delta r^{\overleftarrow{EE}} + q_m(t)(1-q_m(t))\Delta r^{\overleftarrow{EI}}\left(\frac{q(t)}{p(t)}\right) + (q_m(t)-p_m(t))\overline{r}^{\overleftarrow{EI}}\left(\frac{q(t)}{p(t)}\right) \\ \dot{q}_m(t) = & q_m(t)(1-q_m(t))\Delta r^{\overleftarrow{II}} + p_m(t)(1-p_m(t))\Delta r^{\overleftarrow{IE}}\left(\frac{p(t)}{q(t)}\right) + (p_m(t)-q_m(t))\overline{r}^{\overleftarrow{IE}}\left(\frac{p(t)}{q(t)}\right) \end{cases} \tag{S17}$$

To focus on the differentiation between the exposed and the infectious compartments, we study here the variable $Q(t)$ such that:

$$Q(t) = \frac{p_m(t)}{(1-p_m(t))}\frac{(1-q_m(t))}{q_m(t)}. \tag{S18}$$

Thus:

$$\ln(Q(t)) = \mathrm{logit}(p_m(t)) - \mathrm{logit}(q_m(t)).$$

The temporal dynamics of $Q(t)$ is therefore given by:

$$\dot{Q}(t) = \frac{q_m(t)(1-q_m(t))\dot{p}_m(t) - p_m(t)(1-p_m(t))\dot{q}_m(t)}{\Big((1-p_m(t))q_m(t)\Big)^2}.$$

By expanding the expressions for $\dot{p}_m(t)$ and $\dot{q}_m(t)$ and after numerous rearrangements, we obtain:

$$\begin{aligned} \frac{\mathrm{d}\ln(Q(t))}{\mathrm{d}t} = & \underbrace{\frac{q(t)}{p(t)}\left(\frac{1-q_m(t)}{1-p_m(t)}\right)\Delta r^{\overleftarrow{EI}} - \frac{p(t)}{q(t)}\left(\frac{1-p_m(t)}{1-q_m(t)}\right)\Delta r^{\overleftarrow{IE}} + \Delta r^{\overleftarrow{EE}} - \Delta r^{\overleftarrow{II}}}_{\text{Effect of selection } (\mathcal{O}(\varepsilon))} \\ & \underbrace{-\Big(Q(t)-1\Big)\left(\frac{q(t)}{p(t)}\frac{q_m(t)}{p_m(t)}r_m^{\overleftarrow{EI}} + \frac{p(t)}{q(t)}\frac{1-p_m(t)}{1-q_m(t)}r_m^{\overleftarrow{IE}}\right)}_{\text{Effect of "migration" } (\mathcal{O}(1))}. \end{aligned} \tag{S19}$$

In the neutral case ($\varepsilon = 0$), the mutant strain $m$ and the WT strain $w$ have the same phenotype, that is: $\forall (i,j) \in \{E, I\}^2,\ \Delta r^{\overleftarrow{ji}} = 0$, and we rapidly have $p_m(t)/q_m(t) = (1 - q_m(t))/(1 - p_m(t)) = Q(t) = 1$ because "migration" – i.e. transitions between compartments $E$ and $I$, including transmissions – spatially homogenises the frequencies of the variant. Selection will disrupt these quantities to $\mathcal{O}(\varepsilon)$.

Solving (S19) for $Q(t)$ based on a quasi-equilibrium approach, i.e. setting the right-hand sides of (S19) to 0, and using a Taylor expansion for the solution about the neutral case to order $\varepsilon$ yields:

$$Q(t) \approx 1 + \frac{\Delta r^{\overleftarrow{EE}} + \left(\frac{q(t)}{p(t)}\right) \Delta r^{\overleftarrow{EI}} - \left(\frac{p(t)}{q(t)}\right) \Delta r^{\overleftarrow{IE}} - \Delta r^{\overleftarrow{II}}}{\left(\frac{q(t)}{p(t)}\right) r_m^{\overleftarrow{EI}} + \left(\frac{p(t)}{q(t)}\right) r_m^{\overleftarrow{IE}}} + \mathcal{O}(\varepsilon^2). \tag{S20}$$

By replacing the general form of the transition rates with the particular parameters of the model (S1), we get after some rearrangements:

$$Q(t) \approx 1 + \frac{\left(\frac{q(t)}{p(t)}\right)(1 - c(t))\Delta\beta S(t)/N - \left(\frac{1}{q(t)}\right)\Delta\kappa + \Delta\alpha + \Delta\gamma}{\left(\frac{q(t)}{p(t)}\right)(1 - c(t))\beta_m S(t)/N + \left(\frac{p(t)}{q(t)}\right)\kappa_m} + \mathcal{O}(\varepsilon^2). \tag{S21}$$

Note that $q(t)$ and $p(t)/q(t)$ may then be approximated by their quasi-equilibrium values.

Assuming that the virulence may be neglected ($\alpha_m = \alpha_w = 0$) and that there is no difference between the variant and the WT strains in terms of latent period, i.e. $\Delta\kappa = 0$, the previous equation then reduces to:

$$Q(t) \approx 1 + \frac{\left(\frac{q(t)}{p(t)}\right)(1 - c(t))\Delta\beta S(t)/N + \Delta\gamma}{\left(\frac{q(t)}{p(t)}\right)(1 - c(t))\beta_m S(t)/N + \left(\frac{p(t)}{q(t)}\right)\kappa} + \mathcal{O}(\varepsilon^2). \tag{S22}$$

It is interesting to note that the quasi-equilibrium of $Q$ depends on $\Delta\beta$ and $\Delta\gamma$. More specifically, this expression predicts that the value of $Q$ will be greater than 1 in the case of a variant with a higher transmission rate ($\Delta\beta > 0$ and $\Delta\gamma = 0$) while the value of $Q$ will be less than 1 in the case of a variant with a longer duration of infectiousness, i.e. with a lower recovery rate, ($\Delta\gamma < 0$ and $\Delta\beta = 0$). Hence, some data on the differentiation between different host compartments (here between $E$ and $I$) could potentially yield another way to estimate these two quantities.

### S7 Relation with Blanquart *et al.* (2022), eLife

In [1], the growth rate of the epidemic $r(t)$ and the effective reproduction number $\mathcal{R}(t)$ – i.e. the average number of secondary infections – are linked through the framework popularized by Wallinga and Lipsitch in [7]. Let us consider an epidemic that grows exponentially at a rate $r(t)$ and a probability density function $g$ for the generation time – i.e. timing of secondary infections. Assuming that the distribution of the age of infections stabilises very rapidly, the relationship between $r(t)$ and $\mathcal{R}(t)$ are given by [7]:

$$\frac{1}{\mathcal{R}(t)} = \int_0^{+\infty} e^{-r(t)a} g(a)\, \mathrm{d}a. \tag{S23}$$

Let us consider the scenario where a new variant $m$ emerges and spreads in a host population previously dominated by a wild type strain $w$. The selection coefficient associated with the new variant can be computed from the difference in the *per capita* growth rate of the two variants: $s(t) = r_m(t) - r_w(t)$. The higher growth rate of the new variant can be due to different phenotypic effects acting on the transmission and/or the duration of infection and/or the shape of the whole distribution $g$. In our analysis we assume that the mean and the variance of the generation time distribution are linked due to the assumption of exponentially distributed sojourn times. In contrast, other studies have allowed the mean and the variance of the distribution to be independently modified by the mutations of the new variant [1, 6]. More specifically we follow [1] and assume that the variant is characterized by its effective reproduction number $\mathcal{R}_m(t)$ and by its generation time distribution with mean $\mu_m$ and standard deviation $\sigma_m$ (likewise, $\mathcal{R}_w(t)$, $\mu_w$ and $\sigma_w$, respectively, for the resident stain) such that:

$$\begin{cases} \mathcal{R}_m(t) & = \mathcal{R}_w(t)\big(1+\delta_1\big) \\ \mu_m(t) & = \mu_w(t)\big(1+\delta_2\big) \\ \sigma_m(t) & = \sigma_w(t)\big(1+\delta_3\big) \end{cases} \tag{S24}$$

where $\delta_1$, $\delta_2$ and $\delta_3$ refer to the effects of the mutation of the new variant on the three phenotypic traits as in [1]. To characterize these phenotypic differences between the variant and the resident strain, one must then look at $\delta_1$, $\delta_2$ and $\delta_3$.

In the well-known $S(E)IR$ models formalised by a system of ODEs, susceptible hosts $S$ are infected with a constant transmission rate $\beta$ and infectious individuals $I$ recover at a constant rate $\gamma$. In [1], the authors assume that temporal variations in behavior and NPIs would affect the transmission, only captured by variability in the parameter $\mathcal{R}_w(t)$, without affecting the generation time distribution. We use the same assumption in our analysis through $c(t) \in [0;\ 1]$, the effectiveness of NPIs. Accounting for these control measures, the effective reproduction number in classical $S(E)IR$ models is given by: $\mathcal{R}(t) = \frac{(1-c(t))\beta}{\gamma}\frac{S(t)}{N}$, with $S(t)/N$ the proportion of susceptible hosts at time $t$ in the population (of size $N$). We recall the notations made in the main text for the resident strain and the variant, respectively: $\beta_w$ and $\beta_m = \beta_w + \Delta\beta$ referred to transmission rates; $\gamma_w$ and $\gamma_m = \gamma_w + \Delta\gamma$ referred to recovery rates. Thus, assuming $c(t)$ to be the same for both strains, we have:

$$\begin{cases} \mathcal{R}_w(t) & = (1-c(t))\dfrac{\beta_w}{\gamma_w}\dfrac{S(t)}{N} \\ \mathcal{R}_m(t) & = (1-c(t))\left(\dfrac{\beta_w+\Delta\beta}{\gamma_w+\Delta\gamma}\right)\dfrac{S(t)}{N} \end{cases} \tag{S25}$$

As discussed in [6], this is indeed valid for interventions that reduce transmission – e.g. social distancing, face covering – but no longer holds for interventions that lead to isolation of infected individuals – e.g. contact tracing.

In the following, our aim is to show the links between the framework developed in [1] – with phenotypic differences $\delta_1$, $\delta_2$ and $\delta_3$ – and the framework we developed in this study – with phenotypic differences $\Delta\beta$ and $\Delta\gamma$ – through the $SIR$ and $SEIR$ models.

#### S7.1 $SIR$ model

In the classical $SIR$ model formalised by ODEs, the generation time is exponentially distributed (and thus memoryless). Let's start, however, with a gamma-distributed generation time as the exponential distribution is merely a special case of the gamma distribution family. Under this assumption, by substituting $g$ in (S23) for the probability density function of the gamma distribution with mean $\mu_m$ and standard deviation $\sigma_m$, growth rates $r_w(t)$ and $r_m(t)$ thus become [1]:

$$\begin{cases} r_w(t) &= \left(\mathcal{R}_w(t)^{\left(\frac{\sigma_w}{\mu_w}\right)^2} - 1\right)\dfrac{\mu_w}{\sigma_w^2} \\ r_m(t) &= \left(\Big(\mathcal{R}_w(t)\big(1+\delta_1\big)\Big)^{\left(\frac{\sigma_w\left(1+\delta_3\right)}{\mu_w\left(1+\delta_2\right)}\right)^2} - 1\right)\dfrac{\mu_w\big(1+\delta_2\big)}{\Big(\sigma_w\big(1+\delta_3\big)\Big)^2} \end{cases} \tag{S26}$$

Under the assumption of weak selection – i.e. $\delta_1$, $\delta_2$ and $\delta_3$ are small and $\mathcal{O}(\varepsilon)$ – the selection gradient $s(t) = r_m(t) - r_w(t)$ is:

$$\begin{aligned} s(t) = \;& \left(\frac{\mathcal{R}_w(t)^{\left(\frac{\sigma_w}{\mu_w}\right)^2}}{\mu_w}\right)\delta_1 + \left(\left(\mathcal{R}_w(t)^{\left(\frac{\sigma_w}{\mu_w}\right)^2} - 1\right)\frac{\mu_w}{\sigma_w^2} - \frac{2\mathcal{R}_w(t)^{\left(\frac{\sigma_w}{\mu_w}\right)^2}\ln\big(\mathcal{R}_w(t)\big)}{\mu_w}\right)\delta_2 + \\ & 2\left(\frac{\mathcal{R}_w(t)^{\left(\frac{\sigma_w}{\mu_w}\right)^2}\ln\big(\mathcal{R}_w(t)\big)}{\mu_w} - \left(\mathcal{R}_w(t)^{\left(\frac{\sigma_w}{\mu_w}\right)^2} - 1\right)\frac{\mu_w}{\sigma_w^2}\right)\delta_3 + \mathcal{O}(\varepsilon^2). \end{aligned} \tag{S27}$$

At equilibrium (i.e. $\mathcal{R}_w(t) = 1$), $s(t)$ reduces to: $s(t) = \delta_1/\mu_w + \mathcal{O}(\varepsilon^2)$. In **Fig. S13**, we plot an example of relation between $s(t)$ and $\mathcal{R}_w(t)$ according to (S27) for three types of variant. In accordance with [1], we can see that:

- Higher $\delta_1$ are always selected (whatever the value of $\mathcal{R}_w(t)$);
- Lower $\delta_2$ are selected when $\mathcal{R}_w(t) > 1$ (conversely, higher $\delta_2$ are selected when $\mathcal{R}_w(t) < 1$), except in some cases when $\mathcal{R}_w(t)$ becomes too small;
- Higher $\delta_3$ are always selected as soon as $\mathcal{R}_w(t) \neq 1$.

When $\mathcal{R}_w(t)$ is not too far from 1, $\ln\big(\mathcal{R}_w(t)\big) \approx \mathcal{R}_w(t) - 1$, and, eventually assuming that the generation time of the resident strain is exponentially distributed (special case of gamma distribution where $\sigma_w = \mu_w$), (S27) becomes:

$$s(t) \approx \left(\frac{\mathcal{R}_w(t)}{\mu_w}\right)\delta_1 + \left(\frac{\big(\mathcal{R}_w(t)-1\big)\big(1-2\mathcal{R}_w(t)\big)}{\mu_w}\right)\delta_2 + 2\left(\frac{\big(\mathcal{R}_w(t)-1\big)^2}{\mu_w}\right)\delta_3 + \mathcal{O}(\varepsilon^2). \tag{S28}$$

This expression, easier to understand than the previous one, leads to the same interpretations.

We now use the expressions in (S25) for the effective reproduction number of the resident strain and of the variant. Under the assumption of weak selection – i.e. $\Delta\beta$ and $\Delta\gamma$ are small and $\mathcal{O}(\varepsilon)$ –

a Taylor expansion for the effective reproduction number of the variant $\mathcal{R}_m(t)$ about the neutral case ($\varepsilon = 0$) to order $\varepsilon$ yields:

$$\mathcal{R}_m(t) = \mathcal{R}_w(t)\left(1 + \underbrace{\frac{\Delta\beta}{\beta_w} - \mu_w \Delta\gamma}_{\delta_1}\right) + \mathcal{O}(\varepsilon^2), \tag{S29}$$

with $\mu_w = 1/\gamma_w$, the mean generation time for the resident strain.

Likewise, for $\mu_m = 1/(\gamma_w + \Delta\gamma)$, the mean generation time of the variant:

$$\mu_m(t) = \mu_w(t)\left(1 \underbrace{- \mu_w \Delta\gamma}_{\delta_2}\right) + \mathcal{O}(\varepsilon^2), \tag{S30}$$

This result is the same for $\sigma_m$, the standard deviation of the generation time of the variant, as $\sigma_m = \mu_m$ for the exponential distribution. Hence, using the notations of [1]:

$$\begin{cases} \delta_1 &= \dfrac{\Delta\beta}{\beta_w} - \mu_w \Delta\gamma \\ \delta_2 &= -\mu_w \Delta\gamma \\ \delta_3 &= -\mu_w \Delta\gamma \end{cases} \tag{S31}$$

Substituting these expressions for $\delta_1$, $\delta_2$ and $\delta_3$ in (S27) for the exponential case ($\mu_w = \sigma_w = 1/\gamma_w$) along with the expression of $\mathcal{R}_w(t)$ in (S25), $s(t)$ reduces to:

$$s(t) \approx (1 - c(t))\Delta\beta \frac{S(t)}{N} - \Delta\gamma + \mathcal{O}(\varepsilon^2), \tag{S32}$$

which is indeed known to be the expression of the selection gradient in the simplest $SIR$ model [3, 2].

#### S7.2 $SEIR$ model

We now add an exposed state – i.e. infected but not yet infectious –, that individuals leave at a constant rate $\kappa$, altering the generation time distribution. Let us assume that the infectious period is gamma-distributed with mean $\mu^I$ and standard deviation $\sigma^I$ and that the exposed period is exponentially distributed with mean $1/\kappa$. Therefore, the convolution:

$$g(a) = \int_0^a \kappa e^{-\kappa x} \frac{(a-x)^{\left(\frac{\mu^I}{\sigma^I}\right)^2 - 1} \quad e^{-\frac{\mu^I (a-x)}{(\sigma^I)^2}}}{\Gamma\left[\left(\frac{\mu^I}{\sigma^I}\right)^2\right]\left(\frac{(\sigma^I)^2}{\mu^I}\right)^{\left(\frac{\mu^I}{\sigma^I}\right)^2}} \, \mathrm{d}x$$

– where $\Gamma$ is the Gamma function –, yields a probability density function for the generation time with mean $\mu = 1/\kappa + \mu^I$ and standard deviation $\sigma = \sqrt{1/\kappa^2 + (\sigma^I)^2}$. Substituting the probability density function in (S23) for this convolution gives:

$$\mathcal{R}(t) = \left(1 + \frac{r(t)}{\kappa}\right)\left(1 + \frac{\left(\sigma^I\right)^2}{\mu^I} r(t)\right)^{\left(\frac{\mu^I}{\sigma^I}\right)^2}. \tag{S33}$$

The issue with this expression for the $SEIR$ model is that, although it is easy to express $\mathcal{R}(t)$ as a function of $r(t)$, the reverse (expressing $r(t)$ as a function of $\mathcal{R}(t)$) does not seem to be true. Nevertheless, we may look at some special cases.

First, when $1/\kappa \to 0^+$ – i.e. the $SEIR$ model tends to the $SIR$ model since the exposed individuals tend, on average, to leave their compartment instantaneously –, we find indeed the result for the $SIR$ model (S26).

Besides, when the infectious period is now exponentially distributed (with $\sigma^I = \mu^I = 1/\gamma$), the generation time is hypoexponentially distributed (generalized Erlang distribution) and the previous expression becomes:

$$\mathcal{R}(t) = \left(1 + \frac{r(t)}{\kappa}\right)\left(1 + \mu^I r(t)\right), \tag{S34}$$

as already shown in [7], which yields:

$$r(t) = \frac{-\kappa\mu^I - 1 + \sqrt{\left(\kappa\mu^I - 1\right)^2 + 4\kappa\mu^I\mathcal{R}(t)}}{2\mu^I} \tag{S35}$$

It corresponds to the expression we used for the growth rate of the epidemic in this study.

Assuming no change in $\kappa$ between the resident strain $w$ and the variant $m$ – i.e. same latent period, on average, for both strains –, we still have $\mathcal{R}_m(t) = \mathcal{R}_w(t)\big(1+\delta_1\big)$ and we obtain from the expression of the mean generation time $\mu_m$ in (S24) an expression for the mean duration of infectiousness $\mu_m^I$:

$$\begin{aligned} \mu_m(t) = \mu_w(t)\big(1+\delta_2\big) \qquad &\Longleftrightarrow \qquad \frac{1}{\kappa} + \mu_m^I = \left(\frac{1}{\kappa} + \mu_w^I\right)\big(1+\delta_2\big) \\ &\Longleftrightarrow \qquad \mu_m^I = \mu_w^I + \delta_2\left(\frac{1}{\kappa} + \mu_w^I\right) \end{aligned} \tag{S36}$$

Substituting $\mathcal{R}(t)$ and $\mu^I$ in (S35) using (S24) and (S36), respectively, we can calculate the selection gradient $s(t) = r_m(t) - r_w(t)$. Again, a Taylor expansion about the neutral case (weak selection) gives:

$$\begin{aligned} s(t) = \; & \left(\frac{\kappa\mathcal{R}_w(t)}{\sqrt{\left(\kappa\mu_w^I - 1\right)^2 + 4\kappa\mu_w^I\mathcal{R}_w(t)}}\right)\delta_1 + \\ & \left(\frac{\left(\kappa\mu_w^I + 1\right)\left(\sqrt{\left(\kappa\mu_w^I - 1\right)^2 + 4\kappa\mu_w^I\mathcal{R}_w(t)} + \kappa\mu_w^I\Big(1 - 2\mathcal{R}_w(t)\Big) - 1\right)}{2\kappa\left(\mu_w^I\right)^2\sqrt{\left(\kappa\mu_w^I - 1\right)^2 + 4\kappa\mu_w^I\mathcal{R}_w(t)}}\right)\delta_2 + \mathcal{O}(\varepsilon^2). \end{aligned} \tag{S37}$$

At equilibrium (i.e. $\mathcal{R}_w(t) = 1$), $s(t)$ is simply: $s(t) = \delta_1 / \left(1/\kappa + \mu_w^I\right) + \mathcal{O}(\varepsilon^2)$. Furthermore, in any case, we also have:

- Higher $\delta_1$ are always selected (whatever the value of $\mathcal{R}_w(t)$);
- Lower $\delta_2$ are selected when $\mathcal{R}_w(t) > 1$ (conversely, higher $\delta_2$ are selected when $\mathcal{R}_w(t) < 1$).

As in the previous subsection with the $SIR$ model, a weak selection approximation of $\mathcal{R}_m(t)$ from (S25) yields:

$$\mathcal{R}_m(t) = \mathcal{R}_w(t)\left(1 + \underbrace{\frac{\Delta\beta}{\beta_w} - \mu_w^I \Delta\gamma}_{\delta_1}\right) + \mathcal{O}(\varepsilon^2), \tag{S38}$$

and, for the mean generation time of the variant $\mu_m = 1/\kappa + 1/(\gamma_w + \Delta\gamma)$:

$$\mu_m(t) = \mu_w(t)\left(1 \underbrace{- \frac{\kappa\left(\mu_w^I\right)^2}{\kappa\mu_w^I + 1}\Delta\gamma}_{\delta_2}\right) + \mathcal{O}(\varepsilon^2). \tag{S39}$$

Hence, with the notations of [1]:

$$\begin{cases} \delta_1 & = \dfrac{\Delta\beta}{\beta_w} - \mu_w^I \Delta\gamma \\ \delta_2 & = -\dfrac{\kappa\left(\mu_w^I\right)^2}{\kappa\mu_w^I + 1}\Delta\gamma \end{cases} \tag{S40}$$

Substituting these expressions for $\delta_1$ and $\delta_2$ in (S37) along with the expression of $\mathcal{R}_w(t)$ in (S25) and $\mu_w^I = 1/\gamma_w$, the selection gradient becomes after some rearrangements:

$$\begin{aligned} s(t) \quad = \quad & \left(\frac{\kappa}{\sqrt{\left(\kappa - \gamma_w\right)^2 + 4\kappa(1 - c(t))\beta_w \frac{S(t)}{N}}}\right)(1 - c(t))\Delta\beta\frac{S(t)}{N} \\ & - \frac{1}{2}\left(\frac{\kappa - \gamma_w}{\sqrt{\left(\kappa - \gamma_w\right)^2 + 4\kappa(1 - c(t))\beta_w \frac{S(t)}{N}}} + 1\right)\Delta\gamma + \mathcal{O}(\varepsilon^2). \end{aligned} \tag{S41}$$

This is the theoretical derivation of the selection gradient we used in this study (cf. equation (S14)).
